# The diagnostic and prognostic utility of mitral annular plane systolic excursion (MAPSE): A systematic review

**DOI:** 10.1101/2025.03.05.25323051

**Authors:** Debbie Falconer, Fredrika Fröjdh, Nikolas Wyeth, Daniel Brieger, Gaby Captur, Rebecca Kozor, Martin Ugander

## Abstract

Movement of the mitral annulus towards the left ventricular (LV) apex during systole, termed atrioventricular plane displacement (AVPD) or mitral annular plane systolic excursion (MAPSE), was first observed by Leonardo da Vinci in the 15^th^ century. MAPSE, a measure of longitudinal movement, shows good agreement between transthoracic echocardiography and cardiac magnetic resonance imaging (CMR), and can also be measured by transesophageal echocardiography and gated cardiac computed tomography. Manual measurement is fast, simple, and less reliant on good echocardiographic image quality than left ventricular ejection fraction (LVEF) or global longitudinal strain (GLS). Also, measurement can be easily automated, reducing reporting time. However, no major imaging guidelines advise routine measurement. We present a systematic review of studies appraising the diagnostic and prognostic performance of MAPSE from PubMed, Medline, Google Scholar and Embase until September 2025 in accordance with the PRISMA statement. Our findings demonstrate that MAPSE correlates with both LVEF (*r*=0.64 [95% confidence interval 0.54– 0.74]) and GLS (*r*=0.53 [0.45–0.63]), thus showing a modest association with measures of systolic function that may be particularly useful in patients with poor echocardiographic windows. Importantly, MAPSE falls while LVEF remains preserved across a range of pathologies, enabling earlier detection of systolic impairment than when using LVEF. MAPSE is also a powerful prognostic tool, outperforming both LVEF and GLS in predicting adverse events in several studies. Taken together, MAPSE has a clinically useful and important role that merits integration into routine cardiac imaging and care.

## Introduction

First described by Leonardo Da Vinci in the 15^th^ Century (1), mitral annular plane systolic excursion (MAPSE) refers to the movement of the mitral annulus towards the apex during systole arising from ventricular contraction (2). MAPSE has also been referred to as movement of the mitral ring or atrioventricular plane displacement (AVPD). The term MAPSE came into common use in the early 2000s, and for simplicity, shall be used throughout this review. In the majority of patients, the cardiac apex is effectively fixed relative to the chest wall meaning long-axis contraction of the heart can be measured through the change in position of the mitral annulus. MAPSE, therefore, is a marker of longitudinal left ventricular (LV) movement, which contributes 60% of the total stroke volume (3). This proportion is constant across healthy hearts, elite athletes, and patients with dilated (4) and ischaemic cardiomyopathy, even when the absolute longitudinal function is decreased (5).

Despite a growing body of literature pointing to the usefulness of MAPSE as a measure of cardiac function, the majority of contemporary imaging protocols do not advise routine measurement, including the most recent guidelines from the British Society for Echocardiography (6) (BSE), American Society of Echocardiography (ASE) (7), European Association of Cardiovascular Imaging (EACVI) (8), and the Society for Cardiovascular Magnetic Resonance (SCMR) (9). Although the BSE recommends a single MAPSE measurement as part of a comprehensive dataset (6), it is not routinely performed in most institutions. EACVI suggests MAPSE can be measured if other markers of longitudinal function are not available (8).

Left ventricular ejection fraction (LVEF) is the mainstay for assessing systolic function, and normal LVEF can be maintained until late stages of a disease by a compensatory increase in radial function (10,11), meaning early systolic impairment is missed when assessed by LVEF alone. Global longitudinal strain (GLS) is a myocardial deformation analysis reflecting primarily longitudinal contraction, and it detects changes in systolic function before LVEF is impaired (12). Strain can be expressed as a function of the entire LV, termed long-axis strain, or on a regional basis, the latter of which has been widely adopted in cardio-oncology. Both GLS and LVEF are highly dependent on image quality. Conversely, MAPSE requires only measurement of mitral annular movement, so is less limited by imaging quality or user experience (13). MAPSE can be measured using transthoracic echocardiography (TTE) (**Figure 1**), cardiac magnetic resonance (CMR) (**Figure 2**), cardiac computer tomography (CT), and transesophageal echocardiography, and shows good agreement between modalities (14). MAPSE can also be easily automated, reducing reporting time (15). Therefore, excluding MAPSE from routine imaging may result in the loss of an easily attainable but informative imaging biomarker.

**Figure 1.**
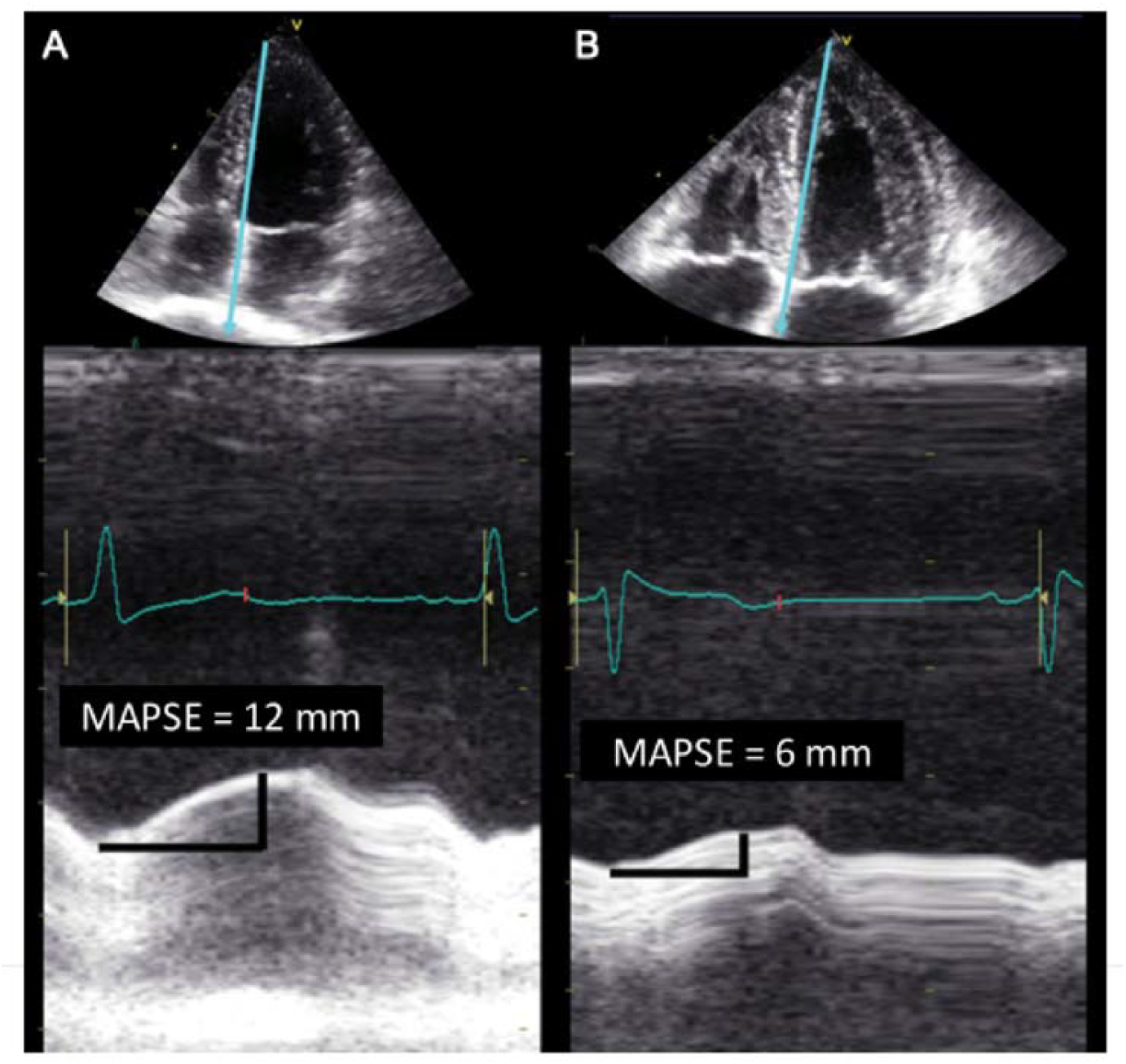
Examples of MAPSE measurement using M-mode echocardiograph in a person with (A) normal systolic function and (b) impaired systolic function. Reproduced with permission from Hu et al^13^, 2012. **Abbreviation:** MAPSE, mitral annular plane systolic excursion

**Figure 2.**
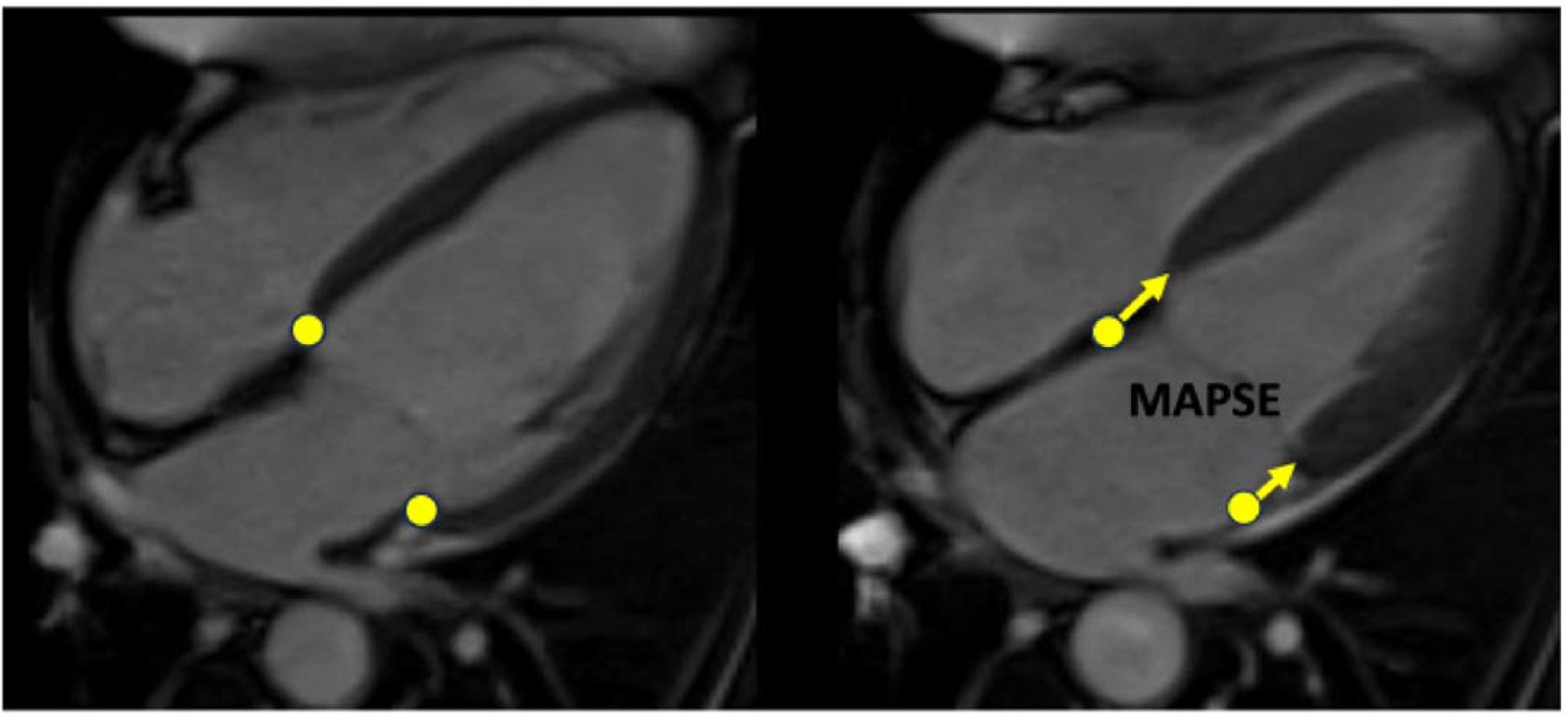
Example of MAPSE measurement using CMR. MAPSE measurement using CMR on 4-chamber cine view from lateral and septal annulus. Adapted with permission from Mayr et al^89^, 2020. Abbreviations as per **Figure 1**.

This study aimed to systematically review evidence outlining the diagnostic and prognostic utility of MAPSE in adult cardiology patients, where possible comparing its performance to LVEF and GLS. The review addresses the following clinical questions:

1. What are the normal values for MAPSE?
2. When LVEF and GLS are not available, can MAPSE be used to diagnose systolic dysfunction (i.e. how closely does MAPSE correlate to these measurements)?
3. For what pathologies can MAPSE be used to detect early systolic dysfunction in the presence of a normal LVEF?
4. In cardiovascular disease, how strongly is MAPSE associated with prognosis, and how does this compare with LVEF and GLS?

## Methods

This systematic review is presented in accordance with the PRISMA statement (16). PubMed, Medline, Embase and Google Scholar were searched (terms in **supplementary material**). Retrospective and prospective observational studies assessing the diagnostic and prognostic utility of MAPSE in adult humans until September 2025 were included. Studies of healthy cohorts and those with pathology were included, and were grouped into the following categories:

1. Studies defining normal values
2. Diagnostic studies:
  a. Measuring association between MAPSE with other markers of systolic function.
  b. Comparing MAPSE in populations with a known pathology vs healthy controls where LVEF is normal.
3. Prognostic studies measuring the association between MAPSE and outcomes in cardiovascular disease and comparison with LVEF and GLS where available.

Non-English publications, abstracts and studies pertaining to congenital heart disease were excluded (**Supplementary Table S-1**). Review articles were not included, but references were manually searched for additional publications. Titles and abstracts were independently screened by both reviewers (DF and FF). If abstracts were insufficient to determine whether the publication should be included or not, the full text was read to inform the final decision and in cases of uncertainty a third reviewer (MU) was consulted. Data from each study was extracted and tabulated by one reviewer (DF) and confirmed by another reviewer (FF). Collected data included study author and year, number of participants, pathology, imaging modality, measurement point, and outcomes. Outcomes included correlations and MAPSE values per group for diagnostic studies, and hazard ratios or odds ratios for prognostic studies as reported.

Meta-analysis of correlation coefficients (*r*) was calculated as a mean correlation [95% confidence interval] weighted by sample size to analyse overall correlation between MAPSE and LVEF or GLS using the Hunter-Schmidt method (17) in R (PsychMeta package, R version 4.2.3, 2023). Meta-correlation was calculated for each measurement site (e.g. lateral MAPSE) when reported by at least two studies. High agreement has previously been reported between CMR and TTE measurements (14), and between manual and automated approaches (15), therefore these modalities were considered sufficiently comparable to be analysed together.

For studies assessing MAPSE as a diagnostic marker when LVEF is normal and as a prognostic marker, there was substantial heterogeneity in measurement location, disease populations, and diagnostic/ prognostic criteria. Therefore, a formal meta-analysis was deemed inappropriate and the results are described narratively.

## Quality Assessment

The methodological quality of included cohort and cross-sectional studies was assessed using the Newcastle-Ottawa Scale (NOS) (18) by two reviewers (DF and NW), and a third (MU) was consulted in cases of disagreement. Standard NOS criteria (**Supplementary Table S-2**) was applied to cohort studies reporting diagnostic and prognostic outcomes. For cross-sectional studies reporting MAPSE at a single timepoint in groups with preserved LVEF, a modified version of the NOS adapted for cross-sectional studies was used as previously described (19, 20) (**Supplementary Table S-3**). The modified NOS evaluates participant selection, comparability of groups and the assessment of outcomes with a maximum score of 10 (compared with 9 for the standard NOS). Studies were then classified as high (7–9/ 10), moderate (5–6), or low (<5) quality.

As there are no established quality assessment tools specifically designed for studies evaluating correlation between measurements, a formal risk of bias assessment was not conducted for this group of studies.

## Results

The search was performed December 2024–September 2025, and 92 studies were included **(Figure 3)**. Summaries of included studies are presented in **Tables 1–4**.

**Table 1.**
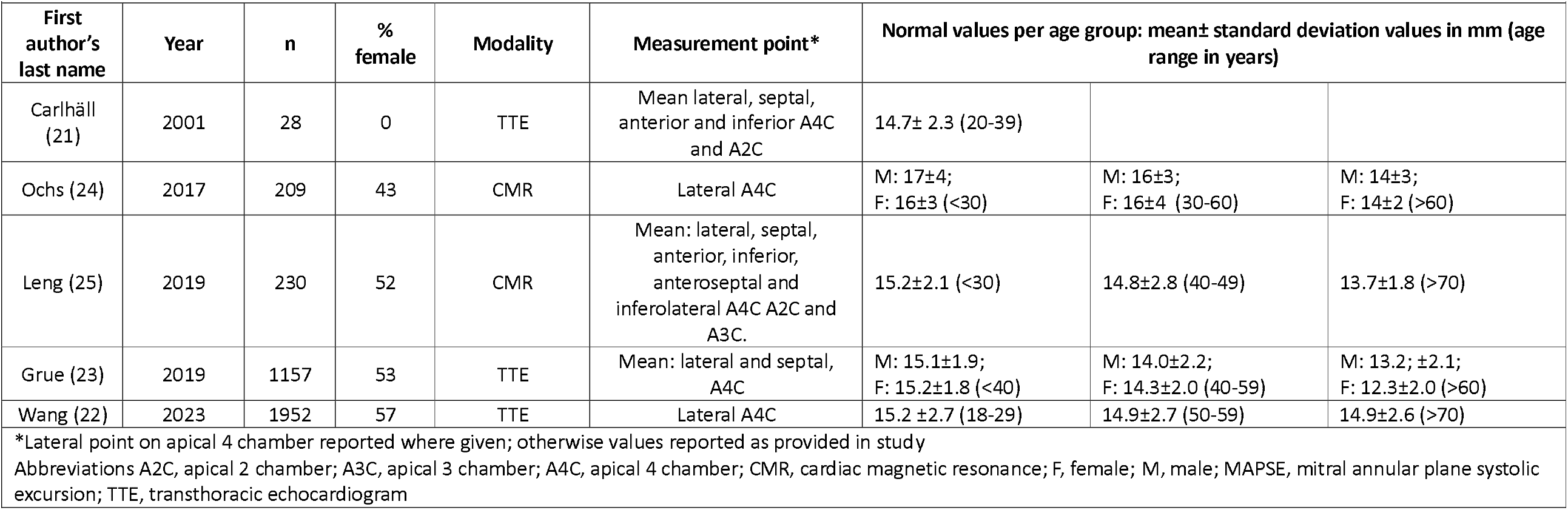
Summary of studies presenting reference values in healthy subjects.

|  |  |  |  |  |  |  |  |  |
| --- | --- | --- | --- | --- | --- | --- | --- | --- |
| First author's last name | Year | n | % female | Modality | Measurement point* | Normal values per age group: mean± standard deviation values in mm (age range in years) |  |  |
| Carlhäll (21) | 2001 | 28 | 0 | TTE | Mean lateral, septal, anterior and inferior A4C and A2C | 14.7± 2.3 (20-39) |  |  |
| Ochs (24) | 2017 | 209 | 43 | CMR | Lateral A4C | M: 17±4;<br>F: 16±3 (<30) | M: 16±3;<br>F: 16±4 (30-60) | M: 14±3;<br>F: 14±2 (>60) |
| Leng (25) | 2019 | 230 | 52 | CMR | Mean: lateral, septal, anterior, inferior, anteroseptal and inferolateral A4C A2C and A3C. | 15.2±2.1 (<30) | 14.8±2.8 (40-49) | 13.7±1.8 (>70) |
| Grue (23) | 2019 | 1157 | 53 | TTE | Mean: lateral and septal, A4C | M: 15.1±1.9;<br>F: 15.2±1.8 (<40) | M: 14.0±2.2;<br>F: 14.3±2.0 (40-59) | M: 13.2; ±2.1;<br>F: 12.3±2.0 (>60) |
| Wang (22) | 2023 | 1952 | 57 | TTE | Lateral A4C | 15.2 ±2.7 (18-29) | 14.9±2.7 (50-59) | 14.9±2.6 (>70) |
| *Lateral point on apical 4 chamber reported where given; otherwise values reported as provided in study<br>Abbreviations A2C, apical 2 chamber; A3C, apical 3 chamber; A4C, apical 4 chamber; CMR, cardiac magnetic resonance; F, female; M, male; MAPSE, mitral annular plane systolic excursion; TTE, transthoracic echocardiogram |  |  |  |  |  |  |  |  |

**Figure 3.**
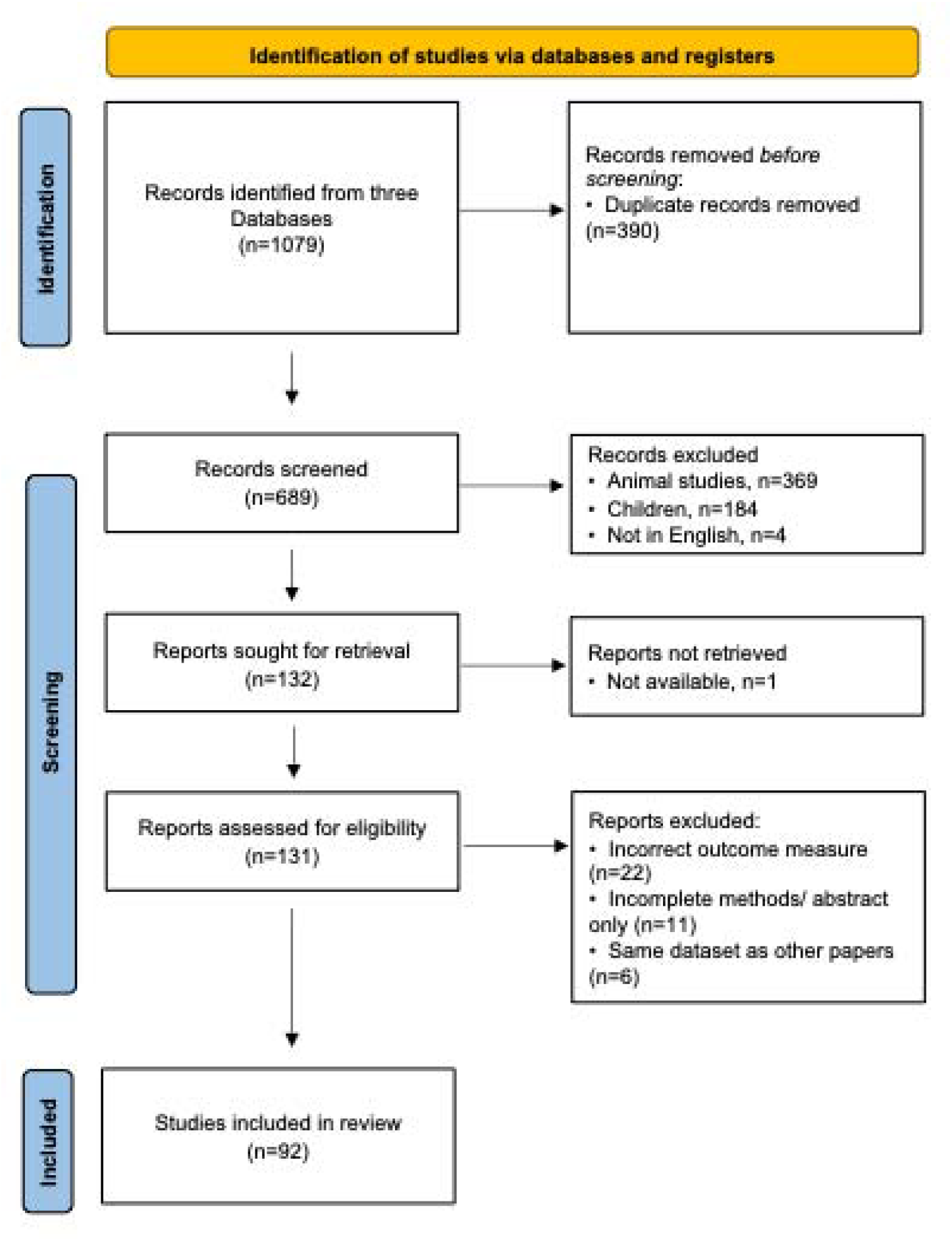
Preferred Reporting Items for Systematic Reviews and Meta-Analyses flow chart outlining study selection. Abbreviations as per **Figure 1**.

### Normal MAPSE values

Five studies (n= 3576) (**Table 1**) report MAPSE values in healthy cohorts. All studies observed an inverse relationship with age (21-25).

### MAPSE as a method of diagnosing systolic impairment

#### 1. Correlation between MAPSE and other markers of systolic function

Limited echocardiographic windows can render accurate assessment of systolic function challenging. The relationship between MASPE and other markers of systolic function has been investigated to ascertain if MAPSE can be used when LVEF or GLS are not available.

### Correlation between MAPSE and LVEF

In a meta-analysis of 21 studies, LVEF and MAPSE demonstrated a moderate pooled correlation across all measurement sites (n = 2864; r = 0.64 ± 0.21 [95% CI: 0.54–0.74]) (21, 26–45); **Table 2; Figure 4**). More specifically, LVEF correlated with MAPSE measured at the lateral apical four-chamber view (n = 2011; r = 0.49 [0.27–0.70]), the mean of lateral and septal sites (n = 968; r = 0.70 [0.45–0.95]), and the mean of lateral, septal, anterior, and inferior sites (n = 366; r = 0.79 [0.56–1.00]).

**Table 2.** Summary of studies comparing MAPSE with other measures of systolic function.

|  |  |  |  |  |  |  |  |  |
| --- | --- | --- | --- | --- | --- | --- | --- | --- |
| First author's last name | Year | Group | N with pathology | N healthy control | Modality | Measurement point with highest correlation | Key findings* |  |
|  |  |  |  |  |  |  | *r* | Optimal threshold if reported |
| Correlation between MAPSE and LVEF |  |  |  |  |  |  |  |  |
| Simonson (26) | 1989 | Mixed cardiac pathologies | 74 | 26 | TTE | Midpoint A4C | 0.84 | <8mm -> LVEF<50%: sens: 98%; spec: 82% |
| Alam (31) | 1991 | Post-ACS | 37 |  | TTE | Mean: lateral, septal, anterior, inferior A2C and A4C (vs radionucleotide angiography LVEF) | 0.87 | ≥10mm -> LVEF≥50% sens 95%; spec 82% |
| Alam (32) | 1992 | Post-ACS or HFrEF | 34 | 45 | TTE | Mean: lateral, septal, anterior, inferior A2C and A4C | 0.96 |  |
| Carlhäll (21) | 2001 | Healthy adults |  | 28 | TTE | Mean: lateral, septal, anterior, inferior A2C and A4C | 0.22 |  |
| Qin (27) | 2004 | Mixed cardiac pathologies | 47 |  | 3D TTE/CMR | Mean: 18 points on mitral annulus (vs CMR LVEF) | 0.84 | <12mm -> LVEF <50% Sens: 95% spec:85% |
| Elnoamany (28) | 2006 | Multiple cardiac pathologies | 133 | 20 | TTE | Mean: lateral, septal, anterior, inferior A2C and A4C | 0.81 | <12mm -> LVEF<50% sens: 90%; spec 88% |
| Bazaz (30) | 2008 | Mixed cohort | 75 |  | TTE | Lateral A4C | 0.87 |  |
| Tsang (29) | 2010 | Multiple cardiac pathologies | 60 |  | TTE/CMR | Midpoint 4AC (vs CMR LVEF) | 0.77 (0.73) |  |
| Matos (39) | 2012 | Mixed cohort | 300 |  | TTE | Mean of lateral and septal A4C | Males 0.64 Females 0.47 | <6mm -> LVEF <30% sens: 92%; spec 85% |
| Zidan (35) | 2016 | Obese ventilated patients | 60 |  | TTE | Mean: lateral, septal, anterior, inferior A2C and A4C | 0.87 |  |
| Lopez-Candales (40) | 2017 | Mixed cohort | 100 |  | TTE | Lateral 4AC | 0.6 |  |
| Magdy (44) | 2018 | Ischaemic cardiomyopathy LVEF<35% | 50 | | Dobutamine stress TTE | Mean: lateral and septal A4C | Baseline 0.28<br>Low dose 0.35 | $\Delta \geq 2$ mm -> contractile reserve >10%<br>sens 68%; spec 59% |
| Hernandez-Burgos (41) | 2018 | Mixed cohort | 150 |  | TTE | Lateral A4C | 0.46 |  |
| Shah (36) | 2019 | Ventilated patients | 39 |  | TTE | Lateral 4AC | 0.61 | >12.5mm -> LVEF >50%<br>sens: 82.1%; spec: 91.7% |
| Stenberg (42) | 2021 | POCUS pre-elective operations | 100 |  | TTE | Mean: lateral and septal A4C | 0.56 | <12mm -> LVEF<50%<br>sens: 95%; spec: 80% |
| Berg (33) | 2022 | HFReF | 287 | 20 | CMR | Mean: lateral, septal, anterior, inferior, anteroseptal and inferolateral A4C A2C and A3C. | 0.65 |  |
| Ozer (43) | 2022 | Consecutive outpatients | 512 |  | TTE | Lateral 4AC | 0.34 |  |
| Bergenzaun (37) | 2013 | ICU admissions septic shock | 50 |  | TTE | Mean: lateral, septal, anterior, inferior A2C and A4C | 0.59 |  |
| Brault (38) | 2023 | Ventilated patients with septic shock | 71 |  | TTE | Lateral A4C | 0.52 |  |
| Turan (34) | 2024 | Duchenne Muscular Dystrophy | 55 | 54 | TTE | Mean: lateral and septal A4C | 0.71 |  |
| Cirin (45) | 2025 | Patients with PPM | 409 |  | TTE | Mean: lateral and septal A4C | 0.89 |  |
| **Correlation between MAPSE and TDI** |  |  |  |  |  |  |  |  |
| Mondillo (47) | 2006 | Healthy subjects | 56 |  | TTE | Mean: lateral, septal, anterior, inferior A2C and A4C | 0.54 |  |
| Ballo (46) | 2008 | Healthy subjects | 92 |  | TTE | Lateral A4C | 0.90 |  |
| **Correlation between MAPSE and GLS** |  |  |  |  |  |  |  |  |
| Luszczak (55) | 2013 | Moderate-severe AS | 82 |  | TTE | Mean: midpoint of A4C, A3C and A2C | −0.56 |  |
| Wenzelburger (53) | 2011 | HFpEF | 62 | 36 | TTE | Mean: lateral and septal A4C | Rest: −0.43<br>Exercise: −0.59 |  |
| Liu (48) | 2014 | HCM | 65 | 48 | TTE (speckle tracking) | Mean: lateral, septal, anterior, inferior, anteroseptal and inferolateral A4C A2C and A3C. | −0.86 |  |
| Chiu (49) | 2016 | Hemodialysis | 198 |  | TTE (speckle tracking) | Mean: midpoint of A4C, A3C and A2C | −0.61 |  |
| Lopez-Candales (56) | 2018 | Mixed cohort | 75 |  | TTE | Lateral A4C | −0.54 |  |
| Teraguchi (54) | 2019 | Moderate-severe MR | 50 |  | TTE (speckle tracking) | Lateral A4C | −0.51 | 14.6% change in lateral MAPSE -> low GLS<br>Sens: 94%; spec: 94% |
| Blixt (50) | 2021 | ICU admissions with septic shock | 73 |  | TTE | Mean: lateral and septal A4C | −0.81 |  |
| Iwakura (51) | 2021 | Post-ACS | 246 |  | TTE | Mean: midpoint A4C and A2C | −0.71 |  |
| Wang (22) | 2023 | Healthy subjects | 1683 |  | TTE | Mid-point A4C (speckle tracking) | −0.35 |  |
| Xue (15) | 2022 | Mixed outpatient cohort | 1572 |  | CMR | Mean: lateral, septal, anterior, inferior, anteroseptal and inferolateral A4C A2C and A3C. | −0.63 |  |
| Berg (33) | 2022 | HFrEF | 287 | 20 | CMR | Mean: lateral, septal, anterior, inferior, anteroseptal and inferolateral A4C A2C and A3C. | −0.76 |  |
| Zhang (57) | 2022 | Chemotherapy-related cardiotoxicity | 200 |  | TTE | Mean: lateral and septal A4C | Baseline: −0.38<br>Post-chemo: −0.53 | Reduction in MAPSE ≥2mm -> 15% drop in GLS<br>Sens: 74.5%; spec 54.9% |
| Brault (38) | 2023 | Ventilated patients with septic shock | 71 |  | TTE | Septal A4C | −0.64 |  |
| Ji (52) | 2024 | Post-heart transplant | 155 | 49 | TTE | Midpoint A4C | -0.71 | <9.1mm associated with mortality/ hospitalisation<br>Sens: 94%; spec 58% |
| Nemes (58) | 2025 | Healthy subjects | 106 |  | TTE | Lateral A4C | -0.35 |  |
| All results are statistically significant with a *p* value <0.05 unless stated otherwise. Where multiple results are provided (e.g. from multiple measurement points) the strongest correlation is given.<br>Abbreviations: A2C, apical 2 chamber; A3C, apical 3 chamber; A4C, apical 4 chamber; ACS, acute coronary syndrome; AUC, area under curve; CABG, coronary artery bypass graft; CMR, cardiac magnetic resonance; LVEF, ejection fraction; HCM, hypertrophic cardiomyopathy; HFpEF, heart failure with preserved ejection fraction; HFrEF, heart failure with reduced ejection fraction; MR, mitral regurgitation; POCUS, point of care ultrasound; sens, sensitivity; spec, specificity; PPM, permanent pacemaker; TDI, tissue Doppler; TOE, transesophageal echocardiography; TTE, transthoracic echocardiogram; VTI, velocity time integral. |  |  |  |  |  |  |  |  |

**Figure 4.**
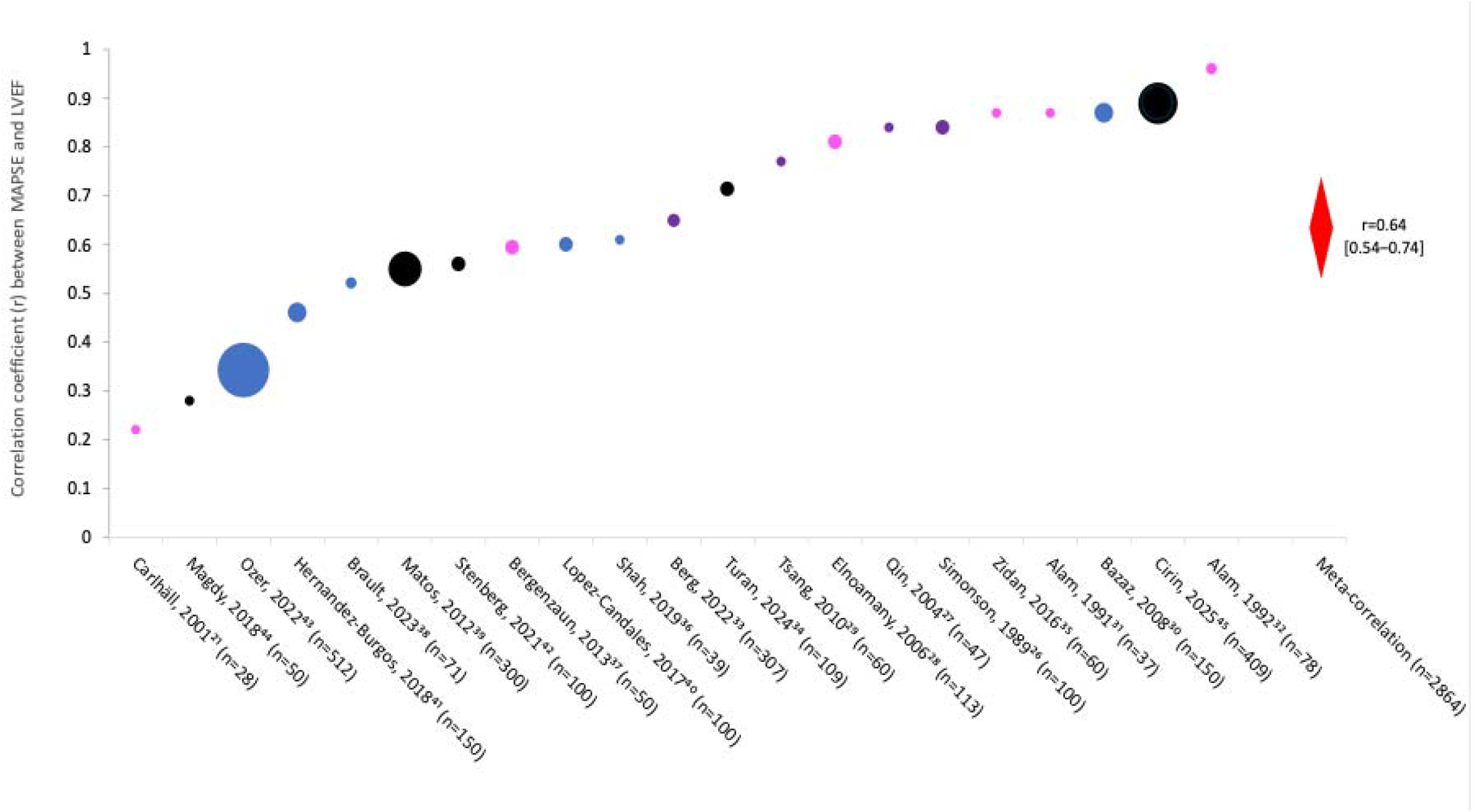
Summary of individual studies and meta-correlation between MAPSE and LVEF. The size of the individual circles denotes the population size. The red diamond denotes the meta-correlation and 95% confidence interval. Studies are colour coded as per measurement site for MAPSE: blue= lateral point on apical 4 chamber view; black= mean of lateral and septal points on apical 4 chamber view; pink= mean of lateral, septal, anterior and inferior points from apical 4 and 2 chamber views; purple= other. Abbreviations: LVEF, ejection fraction, other as per **Figure 1**.

### Correlation between MAPSE and tissue Doppler imaging (TDI)

There was a correlation between MAPSE and tissue Doppler imaging (TDI) in 92 patients (*r*=0.9, p<0.001) (46) and 56 healthy subjects (*r*=0.56, *p*<0.0001) (47). Due to the limited number of studies, no meta-correlation performed was performed for this association.

### Correlation between MAPSE and GLS

A meta-correlation of 15 studies (*n*=5078) showed a moderate pooled correlation between MAPSE and GLS (r=0.53±0.17 [0.45–0.63]) (15,22,33, 38, 48-58) (**Table 2**; **Figure 5**). Specifically, studies using lateral MAPSE alone reported a correlation of 0.45 [0.33–0.73] (n = 231), while studies using the mean of lateral and septal MAPSE reported a stronger correlation of 0.79 [0.56–1.00] (n=366). When MAPSE was measured at six points across three apical views, the correlation was 0.66 [0.46–0.87] (n=1992).

**Figure 5.**
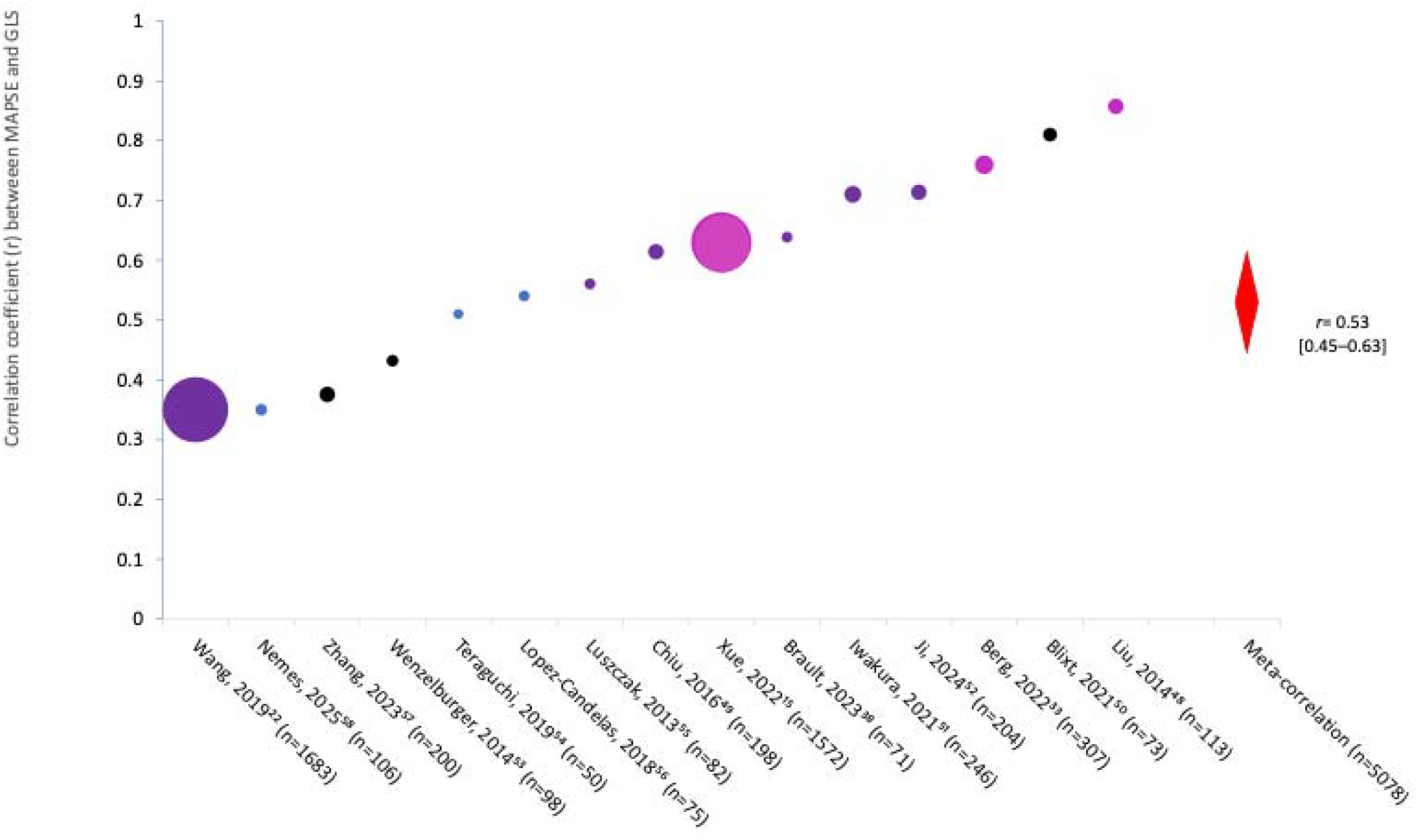
Summary of individual studies and meta-correlation between MAPSE and the absolute value of GLS. The size individual circles denotes the population size. The red diamond denotes the meta-correlation and 95% confidence interval. Studies are colour coded as per measurement site for MAPSE: blue= lateral point on apical 4 chamber view; black= mean of lateral and septal point on apical 4 chamber view; pink= mean of lateral, septal, anterior, inferior, anteroseptal and inferolateral points from apical 4, 2 and 3chamber views; purple= other. **Abbreviations:** GLS, global longitudinal strain; others as per Figure 1.

### Correlation between MAPSE and tissue Doppler imaging (TDI)

There was a correlation between MAPSE and tissue Doppler imaging (TDI) in 92 patients (*r*=0.9, p<0.001) (46) and 56 healthy subjects (*r*=0.56, *p*<0.0001) (47). Due to the limited number of studies, no meta-correlation performed was performed for this association.

### Correlation between MAPSE and GLS

A meta-correlation of 15 studies (*n*=5078) showed a moderate pooled correlation between MAPSE and GLS (r=0.53±0.17 [0.45–0.63]) (15,22,33, 38, 48-58) (**Table 2**; **Figure 5**). Specifically, studies using lateral MAPSE alone reported a correlation of 0.45 [0.33–0.73] (n = 231), while studies using the mean of lateral and septal MAPSE reported a stronger correlation of 0.79 [0.56–1.00] (n=366). When MAPSE was measured at six points across three apical views, the correlation was 0.66 [0.46–0.87] (n=1992).

#### 2. MAPSE as a marker of reduced longitudinal systolic function when LVEF is normal

Thirty-one studies compared MAPSE in patients and controls where LVEF was normal (**Table 3**) (14, 53, 59-79, 81-88). These studies were undertaken in the following patient groups.

**Table 3.** Summary of studies using MAPSE to diagnose left ventricular systolic impairment when LVEF is normal.

|  |  |  |  |  |  |  |  |  |  |
| --- | --- | --- | --- | --- | --- | --- | --- | --- | --- |
| First author's last name | Year | Group | N pathology | N control | Modality | Measurements taken | Key findings |  |  |
|  |  |  |  |  |  |  | Study subgroup (comparison if not healthy controls) | MAPSE pathology (mm) | MAPSE control (mm) |
| Xiao (59) | 2006 | HTN | 26 | 10 | TTE | Septal 4AC |  | 13.1±1.5 | 16.7±2.5 |
|  |  |  |  |  |  | Lateral 4AC |  | 12.3±2.5 | 15.3±0.3 |
| Koulouris (60) | 2005 | HTN | 81 | 50 | TTE | Mean: lateral, septal, anterior, inferior A2C and A4C |  | 10.3±3 | 13.1±1 |
| Ballo (61) | 2010 | DM/HTN | 163 HTN/DM/both | 70 | TTE | Mean: lateral, septal, anterior, inferior A2C and A4C | DM | 13.5±2.3 | 15.1±2.4 |
|  |  |  |  |  |  |  | HTN | 13.2±2.2 |  |
|  |  |  |  |  |  |  | DM+HTN | 11.5±3 |  |
| Salas-Pacheco (62) | 2021 | HTN | 226 | 101 | TTE | Mean: lateral and septal A4C |  | 13.8±2.2 | 15.5±2.0 |
| Tromba (65) | 2017 | Sleeve gastrectomy | 45 |  | TTE | Mean: lateral and septal A4C | Pre- and post-sleeve gastrectomy | Median 16 | Median 18 |
| Li (63) | 2023 | T2DM | 100 | 50 | TTE |  | DM>15 years vs controls | 13.98±1.1 | 15.6±0.45 |
| Loncarevic (64) | 2016 | DM (no HTN) | 210 | 80 | TTE | Mean: lateral, septal, anterior, inferior A2C and A4C |  | 16.4±1.6 | 15.4±2.1 |
| Takeda (67) | 2001 | AS (all grades) | 78 | 10 | TTE | Lateral 4AC | Lower in severe AS vs healthy controls |  |  |
|  |  |  |  |  |  | Septal 4AC | Lower in mild, moderate and severe AS vs control |  |  |
| Rydberg (66) | 2004 | AS (all grades) | 49 | 133 | TTE | Mean: lateral, septal, anterior, inferior A2C and A4C |  | 9.2±2.3 | 10.7±2.8 |
| Weidemann (68) | 2009 | Symptomatic severe AS ± fibrosis on CMR | 21 (no fibrosis) | 21 (severe fibrosis) | TTE | Septal 4AC | Severe fibrosis vs no fibrosis | 4.9±1.0 | 10.7±1.2 |
| Herrmann (71) | 2011 | Low-flow low-gradient AS | 86 |  | TTE | Lateral: and septal A4C | Low-flow low-gradient AS preserved LVEF (vs moderate AS) | 5.8±2.2 | 12.8±1.7 |
|  |  |  |  |  |  |  | Low-flow low-gradient AS reduced LVEF (vs moderate AS) | 5.0±0.9 |  |
| Kempny (69) | 2013 | Severe AS undergoing TAVI | 101 |  | TTE | Lateral A4C | Pre and post-TAVI | 9.1±3.2 | 10.2±3.3 |
| Matos (70) | 2017 | Severe asymptomatic AS | 205 |  | TTE | Mean: lateral, septal, anterior, inferior A2C and A4C | Change in MAPSE requiring intervention scan (vs those not requiring intervention) | 2mm reduction | <2mm reduction |
| Liu (72) | 2018 | Low-flow low-gradient AS | 43 | 20 | TTE | Lateral A4C | Low-flow low-gradient AS reduced LVEF (vs pseudosevere AS) | 9.8±2.6 | 11.6±2.2 |
| Dursunoglu (73) | 2004 | DD | 62 | 40 | TTE | Mean: lateral, septal, anterior, inferior A2C and A4C |  | 12.4±1.4 | 15.3±1.4 |
| Gromadzinski (74) | 2014 | CKD±DD | 36 | 34 | TTE | Lateral A4C | CKD with DD vs CKD without DD median(range) | 13(11–17) | 14(11–16) |
| Wenzelburger (53) | 2011 | HFpEF (during exercise) | 62 | 36 | TTE | Lateral and septal A4C | At rest | 10.9±2.1 | 12.1±2.2 |
|  |  |  |  |  |  |  | During exercise | 12±2.2 | 16.2±2.7 |
| Taşolar (75) | 2015 | High BMI ±DD | 40 high BMI+ HFpEF | 40 high BMI | TTE | Mean: lateral, septal, anterior, inferior A2C and A4C |  | 11.8±1.6 | 15.8±1.6 |
| Bytyci (76) | 2016 | HFpEF | 79 | 29 | TTE | Septal |  | 11.8±2.0 | 14.0±3.0 |
|  |  |  |  |  |  | Lateral |  | 12.4±3.0 | 15.0±3.0 |
| Wierbowska-Drabik (78)) | 2013 | Obesity and LVH | 27 | 27 | TTE | Lateral A4C |  | 20±4 | 22±2 |
| Lopez-Candales (77) | 2014 | Patients with LVH | 45 | 45 | TTE | Lateral A4C |  | 12±2 | 14±2 |
| Qureshi (79) | 2018 | Patients with LVH | 22 | 43 | TTE | Mean: lateral, septal, anterior, inferior A2C and A4C |  | 10.6±0.9 | 13.2±1.7 |
| Doesch (14) | 2015 | HCM | 88 | 50 | TTE/CMR | Mean: lateral and septal A4C |  | 10.0±3.2 | 13.3±3.8 |
| Ozden (81) | 2023 | HCM | 36 | 34 | CMR | Mean: lateral and septal A4C |  | 14.5±2.9 | 11.7±3.2 |
| Kassab (82) | 2025 | Mitral regurgitation | 200 | 50 | CT | Mean midpoint on A3C, A2C and A4C. |  | 9.9±1.3 | 11.9±1.0 |
| Kiotsekoglou (83) | 2008 | Marfan's | 66 | 61 | TTE | Anterior A2C |  | 13.8±0.3 | 18.0±0.2 |
| Harries (84) | 2022 | Anthracycline-related cardiotoxicity | 24 |  | CMR | Lateral A4C | Recovery of function post-anthracycline | 11.7±1.5 (poor recovery) | 15.7±3.1 (good recovery) |
| Botelho (86) | 2024 | Anthracycline-related cardiotoxicity | 5 (with cardiotoxicity) | 56 (without cardiotoxicity) |  |  |  | 13.5 [8.8-16.3] *ns | 13.3 [12.0-13.8] *ns |
| Ichikawa (85) | 2024 | Anthracycline-related cardiotoxicity | 56 (with cardiotoxicity) | 200 (without cardiotoxicity) | TTE | Mean: lateral and septal A4C |  | 14.0±1.5*ns | 13.8±2.6*ns |
| Tafarshiku (87) | 2020 | Hypothyroidism | 81 | 22 | TTE | Lateral A4C |  | 14.9±0.3 | 16.6±0.2 |
| Berger (88) | 2023 | Mixed connective tissue disease | 77 | 59 | TTE | Mean: lateral and septal A4C |  | 13.7±2.1 | 15.3±2.3 |

\* All results presented as mean±standard deviation unless stated; all results are statistically significant with a *p* value <0.05 unless stated otherwise
Written results provided where numerical results not available in manuscripts.
Abbreviations: A2C, apical 2 chamber; A3C, apical 3 chamber; A4C, apical 4 chamber; AS, aortic stenosis; BMI, body mass index; CAD, coronary artery disease; CKD, chronic kidney disease; CT, computed tomography; DD, diastolic dysfunction; DM, diabetes mellitus; LVEF, left ventricular ejection fraction; HFpEF, heart failure with preserved ejection fraction; HFrEF, heart failure with reduced ejection fraction; HTN, hypertension; LGE, late gadolinium enhancement; LVH, left ventricular hypertrophy; ns, not significant; TAVI, transcatheter aortic valve implantation; TTE, transthoracic echocardiogram

#### Hypertension (HTN) or type 2 diabetes mellitus (T2DM)

Six TTE studies of subjects with HTN and/or T2DM demonstrated MAPSE was reduced compared with age and sex-matched controls (59-64). In one study, MAPSE was similar in cohorts with diabetes *or* HTN (13.5±2.3mm and 13.2±2.2mm, respectively) but lower in those with both conditions (11.5±3.0mm, *p*<0.05), suggesting a cumulative effect (61). A study of 45 patients undergoing sleeve gastrectomy showed an increase in MAPSE 6 months post-surgery (median 16mm vs 18m, p<0.01) which may be related to the associated reduction in HTN and DM, though several other cardiovascular risk factors were also reduced (65).

#### Aortic stenosis (AS)

Mean MAPSE was lower in a cohort of 49 patients with AS (all grades) compared with 133 controls (9.2±2.3mm vs 10.7±2.8mm, *p*<0.001) (66). A prospective study (*n*=78) found lateral MAPSE was lower in patients with severe AS compared with controls (*p*<0.002) whilst septal MAPSE was lower in mild, moderate and severe AS (*p*<0.001) (67). In 58 patients undergoing surgical replacement for severe AS, MAPSE was higher in patients with no myocardial fibrosis (10.7±1.2mm) compared with mild (8.8±1.4mm, *p*<0.05) or severe (4.9±1mm, *p*<0.001) changes on biopsy at the time of surgery. Patients with no or mild fibrosis were more likely to experience an improvement in symptoms to New York Heart Association (NYHA) grade I or II, whilst patients with severe fibrosis remained in NYHA class III or IV nine months post-surgery. LVEF did not predict improvement in symptoms (68). Likewise, a study of 101 patients undergoing transcatheter aortic valve implantation reported an increase in MAPSE, but not LVEF, post-procedure (9.1±3.2 to 10.2±3.3mm, *p*=0.006) (69). A study of 205 patients with severe AS showed a reduction in MAPSE of >2mm per serial scan had an increased likelihood of requiring intervention (HR 1.95 [1.04– 3.66] p=0.04) when LVEF remained preserved (70). MAPSE was also lower in low-flow low-gradient severe AS compared with moderate (12.8±1.7mm vs 5.0mm±0.9mm, *p*<0.05) (71) and pseudosevere AS (9.8±2.6 vs 11.6±2.2mm, p<0.05) (72), and may assist in differentiating these groups where there is uncertainty.

#### HfpEF

Five studies found MAPSE is lower in patients with diastolic dysfunction (DD) than controls (53, 73-6). Rather than acting as a marker of pure DD, reduced MAPSE likely reflects concurrent longitudinal systolic dysfunction, as systolic and diastolic function are inextricably linked (13), and may result from a higher prevalence of comorbidities associated with DD such as HTN and DM. In patients with HFpEF undergoing cardiopulmonary exercise testing, MAPSE was lower at rest (10.9±2.1mm vs 12.1±2.2mm, *p*=0.008), and augmented less during exercise in patients with DD compared with controls (12±2.2mm vs 16.2±2.7mm, *p*<0.001) (53).

MAPSE was also lower in three cohorts with LV hypertrophy (LVH) (77-9). In disease states where the LV hypertrophies or dilates, less longitudinal contraction is required to generate the same stroke volume, so a reduction in MAPSE reflects both increased LV mass and size (80)..

#### HCM

Two CMR-based studies (14, 81) found MAPSE was lower in patients with HCM vs controls. Septal MAPSE was reduced compared with lateral MAPSE in patients with outflow obstruction (9.2±3.7mm vs 11±3.6mm, *p*=0.04) reflecting greater impairment in the septum, which is the area of greatest disease (14).

#### Mitral regurgitation

CT-derived MAPSE was lower in patients with severe primary mitral regurgitation (9.9±1.4mm) compared with healthy controls (11.9±1.0mm) and LVEF was preserved relative to their valve function (64±5.5% and 54±6.9% respectively). Furthermore, MAPSE fell as mitral annular calcification increased in severity (8.8±1.0mm vs 5.3±0.7mm for grade 1 and 4 respectively (82).

#### Marfan’s syndrome

MAPSE was lower in all 5 measurement positions in a TTE study compared with controls. Of note, LVEF was also lower in the Marfan’s group, but was well within normal range (66 vs 71%, *p*<0.001) (83).

#### Anthracycline-related cardiac disease (ARCD)

Post-chemotherapy MAPSE was lower in those who did not recover systolic function compared with those with good recovery (11.7±1.5 vs 15.7±3.1mm, *p*=0.03) (84) from ARCD, suggesting MAPSE may identify those most at risk from ongoing impaired function when ARCD occurs. Of note, two studies demonstrated no difference in pre-chemotherapy MAPSE in patients who developed ARCD compared with those who did not (85, 86).

#### Hypothyroidism

A study of 81 patients with hypothyroidism found lateral MAPSE was lower than controls with similar rates of typical cardiovascular risk factors and normal LVEF (17±2 vs 15±3mm, p=0.002), though both groups fell within normal range. (87).

#### Mixed connective tissue disease (MCTD)

MAPSE was lower in 77 patients with MCTD and normal LVEF and GLS compared with controls (88) (13.7±2.1 vs 15.3±2.3, p<0.001).

### MAPSE as a prognostic biomarker

The prognostic capability of MAPSE has been investigated in 29 studies across a range of pathologies (**Table 4**) (15, 37, 43, 51, 52, 89–112).

**Table 4.**
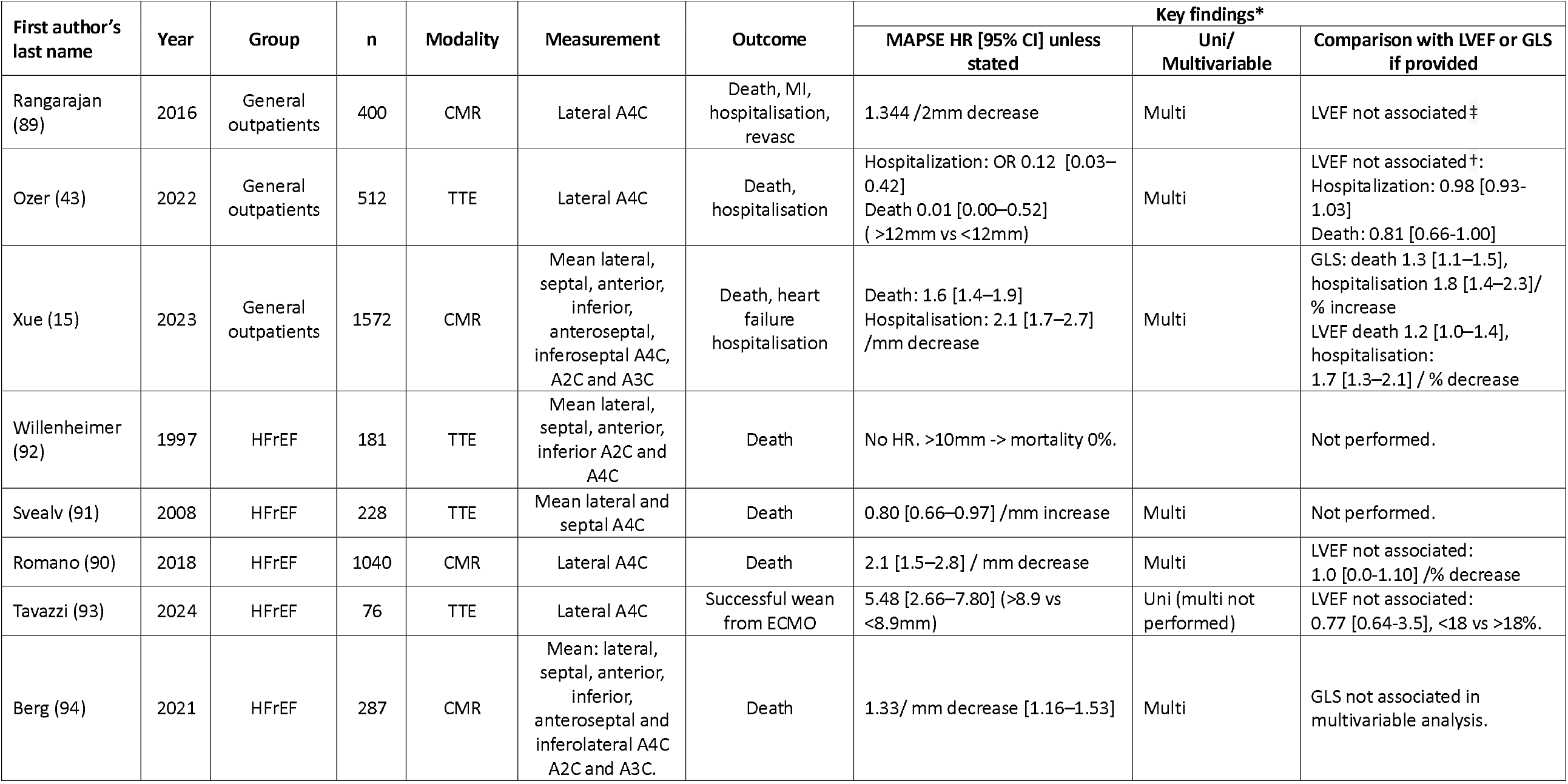
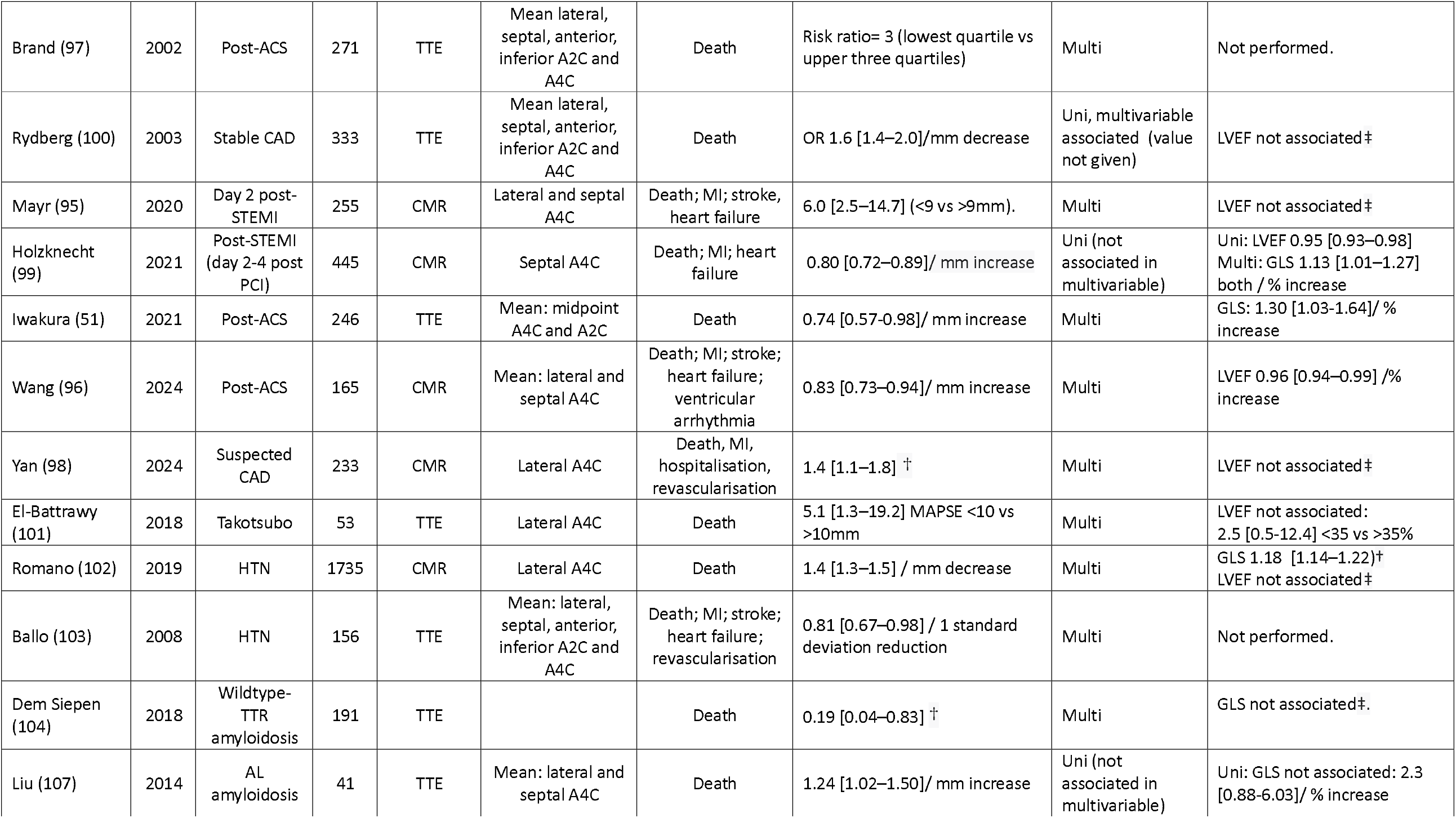
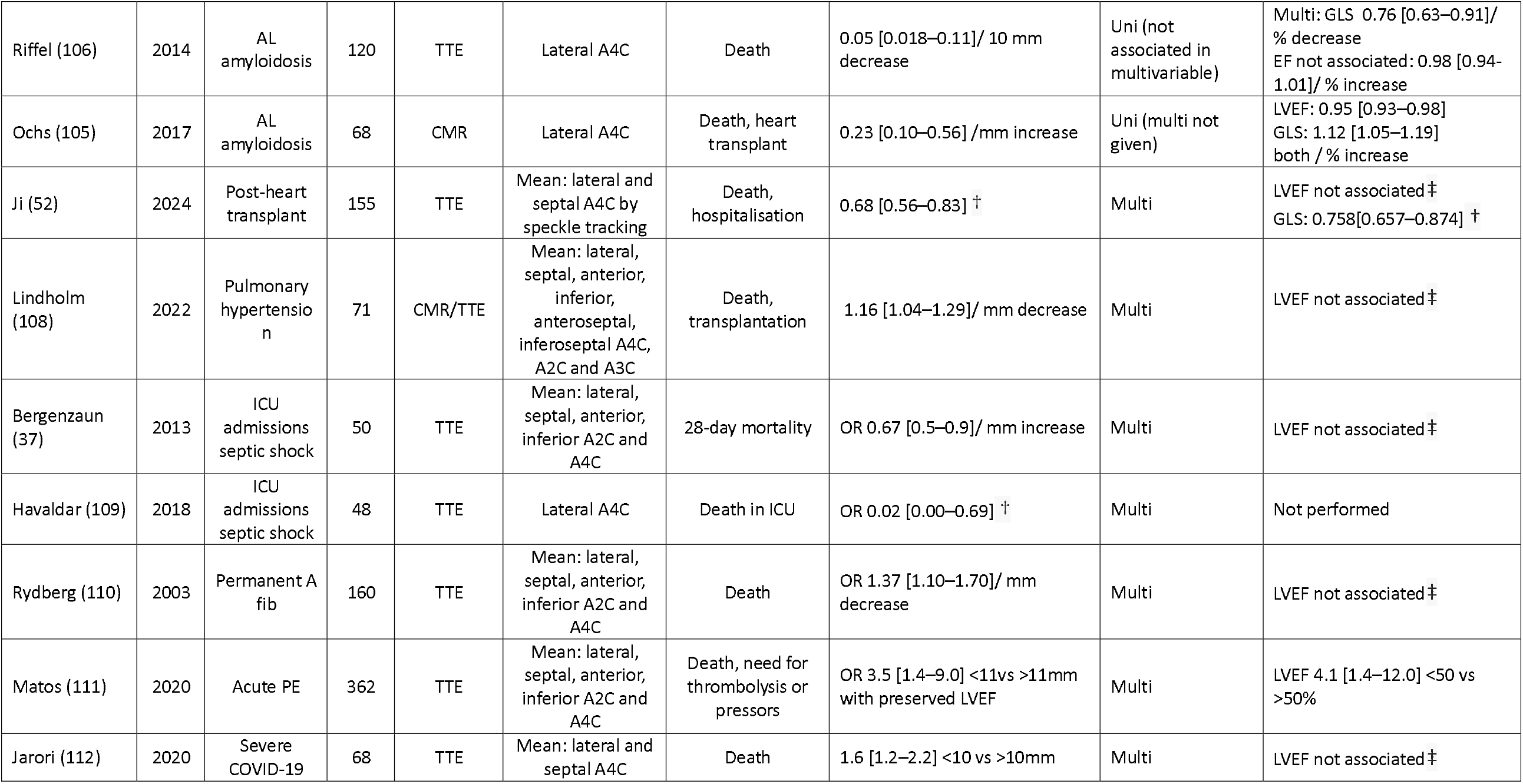
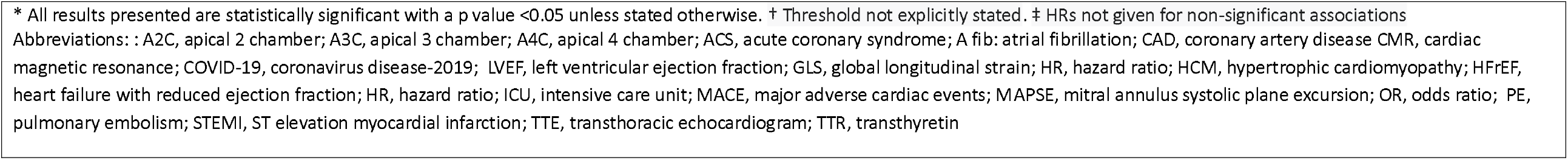
Studies comparing MAPSE with LVEF or GLS as a prognostic marker.

#### General outpatient cohorts

In the largest cohort, which included individuals with pathology and normal scans undergoing outpatient CMR (*n*=1572), automatically-measured MAPSE was a better predictor of death or hospitalisation for heart failure (HR 1.7 [1.5–2.0] per mm decrease, *p*<0.001) compared with GLS (HR 1.5 [1.3–1.7] per % increase, *p*<0.001) or LVEF (HR 1.4 [1.2–1.6] per % decrease, *p*<0.001) on multivariable analysis (15). A similarly heterogeneous cohort of 400 patients followed up for 14 months, found lateral MAPSE was the strongest imaging-based predictor of MACE after multivariable analysis (HR 1.34 per 2mm decrease, *p*=0.02) whereas neither LVEF nor the presence of LGE remained associated (89). In a TTE-based study of 512 patients, lateral MAPSE was the only imaging-based predictor of mortality (HR 0.01 [0.00–0.52] for MAPSE ≥12 vs <12 mm, *p*=0.02) (43), as early as 30 days after imaging (*p*<0.001).

#### HfrEF

Multiple CMR and TTE studies have demonstrated the prognostic significance of MAPSE (90-94). The largest, a multicentre study of 1040 patients with HFrEF undergoing CMR showed lateral MAPSE was the strongest imaging-based biomarker in predicting mortality (HR 2.1 [1.5– 2.8] per mm decrease, *p*<0.001). Presence of LGE was a weaker predictor (HR 2.0 [1.3–3.0], *p*=0.02) whilst LVEF was not associated. Patients with median MAPSE >9mm had a similar survival regardless of LVEF (*p*=0.96) (90). When compared, MAPSE outperformed LVEF (90,93) and GLS (94) in predicting prognosis.

#### IHD

CMR-measured MAPSE was a better predictor of major adverse cardiac events (MACE) (death, MI, stroke, heart failure) than LVEF in an ST-elevation myocardial infarction (STEMI) cohort followed up for 3 years (area under curve 0.74 vs 0.61, *p*<0.01) (95). MAPSE <9mm remained a predictor of MACE in models of clinical and imaging risk factors (HR 6.0 [2.5– 14.7], *p*<0.01) where LVEF was not associated. Another study of 165 patients post-acute coronary syndrome (ACS) also found MAPSE was a stronger predictor of MACE than LVEF (HR 0.83 [0.73–0.94] per mm increase, *p*=0.004 vs HR 0.96 [0.94– 0.99] per % decrease, *p*=0.01) (96). In 271 patients with suspected ACS, those with MAPSE <8mm had a risk ratio of 3.0 (*p*<0.001) for death or heart failure hospitalisation during follow-up compared with the remainder of the cohort (97). In a different cohort with suspected ACS (n=233) lateral MAPSE on CMR predicted MACE during follow up (HR 1.4 [1.1– 1.8]) (95), whereas LVEF did not. Conversely, another CMR study of 445 patients 2-4 days post-revascularisation for STEMI found MAPSE was a predictor in univariable analysis (HR 0.80 [0.72–0.89] per mm increase), but not when adjusted for other imaging biomarkers (99). This difference may arise from the model’s use of CMR-biomarkers such as infarct size or microvascular obstruction that were not adjusted for in other studies.

In 333 patients with stable IHD, MAPSE was the only imaging marker that correlated independently with mortality (OR 1.6 [1.4–2.0] per mm decrease, *p*<0.001) (100).

#### Takotsubo cardiomyopathy

Among patients (n=53) with MAPSE <10mm on admission, mortality was higher after 12 months follow-up, but there was no difference within the first year. MAPSE <10mm was a predictor of mortality (HR 5.1 [1.3–19.2], *p*<0.01), whilst LVEF <35% was not (101).

#### Hypertension

5.1 years following a CMR (n=1735) lateral MAPSE was associated with increased mortality (HR 1.4 [1.3–1.5] per mm decrease, *p*<0.001) in the whole cohort, as well as in the subgroup analysis of patients with preserved LVEF (HR 1.3 [1.2–1.5] per mm decrease, *p*<0.001) (102). GLS was a predictor of MACE in multivariable analysis, but weaker than MAPSE (HR 1.18 [1.14–1.22]per % increase, *p*<0.001) and LVEF was not a predictor at all. A TTE study (n=156) also reported mean MAPSE was a predictor of MACE in multivariable analysis (HR 0.81 [0.67– 0.98 per standard deviation reduction, *p*<0.05) (103).

#### HCM

MAPSE was lower in those with MACE (death, transplant or ventricular arrhythmia) (6.9±1.9mm vs 10.2±3.1mm, *p*=0.01) with no difference in LVEF between groups (*p*=0.86) (14).

#### Amyloidosis

In 191 patients with wildtype transthyretin amyloidosis, MAPSE was the only imaging-based predictor of mortality in multivariable analysis (HR 0.19 [0.04–0.83], *p*=0.003) (104). A CMR-based study of 68 AL amyloid patients showed lateral MAPSE was a stronger predictor of death or heart transplant (HR 0.23 [0.10–0.56] per mm decrease, *p*=0.001) than LVEF or GLS (105). However, two other TTE studies of patients with AL amyloid found MAPSE predicted death in uni-, but not multivariable analysis (106, 107). MAPSE was a stronger predictor than LVEF in both cohorts, but did not out-perform GLS.

#### Heart transplant

MAPSE measured by STE at the midpoint of the mitral annulus in 155 patients was a stronger predictor of death/hospitalisation than GLS (HR 0.68 [0.56– 0.83] vs 0.76 [0.66–0.87], *p*<0.001) or LVEF (not associated) (52). Moreover, MAPSE was obtainable in a greater proportion (86%) compared to LVEF or GLS (72%) (52).

#### Pulmonary arterial hypertension (PAH)

In 71 patients with PAH, MAPSE by CMR was higher in survivors (12.3±3.0mm vs 10.0±3.0mm, *p*=0.02). MAPSE predicted death or transplantation (HR 1.2 [1.0– 1.3] per mm decrease *p*=0.01) whilst LVEF and right ventricular EF were not associated (108).

##### Septic shock

Two studies found MAPSE to be the sole or strongest imaging biomarker for predicting mortality in multivariable analysis (37,109). LVEF was not a predictor of mortality in any study.

#### Atrial fibrillation (AF)

A study of 160 patients with AF showed MAPSE was a predictor of mortality (OR 1.4 [1.1–1.7] per mm decrease, p<0.01) where LVEF was not. (110).

##### Acute pulmonary embolism (PE)

MAPSE <11mm predicted mortality or the need for advanced therapies in 363 patients with acute PE and preserved LVEF (OR 3.5 [1.4–9.0], *p*=0.01). MAPSE <11mm combined with tricuspid annular plane systolic excursion (TAPSE) <16mm was strongly associated with adverse outcomes (OR 10.8 [3.1–37.8], *p*<0.01). LVEF <50% was also an adverse prognostic marker (OR 4.1 [1.4–12.0], *p*=0.011) (111).

#### Coronavirus disease-2019 (COVID-19)

MAPSE was the only left-heart imaging biomarker that differed in patients who died and those who survived (11±3 vs 12±3mm, *p*=0.006) (112). In multivariable analysis, MAPSE (HR 1.6 [1.2– 2.2] per mm decrease, *p*=0.006) and right atrial volume index (RAVI) were predictors of mortality. Difference in RAVI is likely to represent the higher ventilatory requirements of the more unwell patients.

### Quality of evidence assessment

Overall, the studies demonstrated high methodological quality. NOS and modified-NOS scores ranged from 6–9, and all-but-one study was rated as high quality (7 or higher). No studies were rated as low-quality. A full breakdown of individual study scores is provided in **Supplementary Tables S-4–6**.

## Discussion

This systematic review found a moderate-to-strong correlation between MAPSE and LVEF, and a moderate correlation between MAPSE and GLS. In cohorts with a normal LVEF, MAPSE was lower in patients with HTN, T2DM, AS, HCM, Marfan’s syndrome, hypothyroidism and mixed connective tissue disease compared with matched healthy controls. Measuring MAPSE routinely, and monitoring its change over time in these conditions, may allow detection of early systolic impairment that would otherwise be missed.

In the largest study of general outpatients, MAPSE was the strongest predictor of death or hospitalisation, outperforming LVEF and GLS (15). MAPSE predicted mortality or MACE in cohorts with HFrEF, IHD, Takotsubo cardiomyopathy, HTN, amyloidosis, post-heart transplant, pulmonary hypertension, AF and COVID-19, where LVEF was either a weaker predictor or not associated. MAPSE was also a stronger predictor than GLS in cohorts with hypertension and amyloidosis. Taken together, routine incorporation of MAPSE into clinical studies with any of the above conditions would seem appropriate to identify the most at-risk patients.

A specific clinical context in which MAPSE may provide additional value for guiding treatment is aortic stenosis. In this setting, MAPSE — but not LVEF — has been shown to improve following intervention (69) and is associated with postoperative symptomatic improvement (68). Yet, current guidelines recommend reduced LVEF as an indication for intervention in asymptomatic patients (113). Importantly, long-axis movement can also be assessed using gated cardiac computed tomography (82), which is routinely performed as part of the pre-intervention workup.

### Measurement considerations

In this study, the strongest correlation with LVEF was observed using the mean MAPSE derived from lateral, septal, anterior, and inferior sites. The strongest correlation with GLS was found using the mean of lateral and septal MAPSE. Although incorporating additional measurement sites may require more operator-time, our data suggests it may provide greater diagnostic value and a more comprehensive assessment of systolic function. Automated measurements have proven possible using both TTE and CMR, and show good agreement with manual measurements (114). This may be of value particularly in patients with atrial fibrillation, where MAPSE is likely to vary from beat to beat and may require an average of multiple measurements to be taken. MAPSE can also be measured by speckle tracking echocardiography, which automatically tracks and measures annular movement with less angle dependence, but is only available with specific vendor software.

### Limitations of MAPSE

The main limitation with MAPSE is the lack of regional assessment of LV function (115), for which visual assessment and strain remain essential. As the use of M-mode on TTE is angle dependent, off-axis views may underestimate MAPSE, reducing specificity. However, MAPSE can readily be measured in a less angle-dependent fashion using a simple linear caliper applied to a 2D image. MAPSE is also reduced in patients having undergone mitral valve replacement (116), when it may reflect post-surgical changes moreso than inherent LV myocardial function. Finally, as mentioned, widespread application of MAPSE is currently limited by a lack of standardised practice. However, the generation of normal values from large datasets may lead to the inclusion of MAPSE in guidelines, and with it set the standards for acquisition, measurement, and interpretation.

### Limitations of this study

Study quality was assessed using the NOS, which, while widely used, is subjective and may not fully capture all sources of bias in observational studies. As aforementioned, there was significant methodological heterogeneity between studies, particularly in the measurement sites and, as well as study populations and outcome definitions. These variations limited the ability to perform a formal quantitative meta-analysis and precluded direct pooling of results. As a result, it is not possible to derive firm numerical thresholds to guide clinical practice currently. Instead, our findings underscore the promising clinical potential of MAPSE.

## Conclusions

MAPSE is a marker of global longitudinal LV function and has a strong association with clinical prognosis. Measurement of MAPSE allows assessment of LV systolic function even with compromised echocardiographic image quality. Early systolic impairment that may be missed when using LVEF alone can be detected using MAPSE. MAPSE also aids in prognostication across multiple cardiac conditions including hypertension, aortic stenosis, and ischaemic heart disease, outperforming LVEF and GLS in several pathologies. Despite the advancement of more sophisticated techniques within cardiac imaging, new studies continue to highlight the clinical information that MAPSE has to offer clinicians and patients. Those contemplating using MAPSE in their clinical practice may consider both the wealth of data supporting its use presented herein, and da Vinci’s timeless sage words that “simplicity is the ultimate sophistication” (117).

## Data Availability

All data produced in the present work are contained in the manuscript

## Abbreviations and acronyms

ACS: acute coronary syndrome
AVPD: atrioventricular plane displacement
AS: aortic stenosis
AF: atrial fibrillation
AI: artificial intelligence
ARCD: anthracycline related cardiac dysfunction
BSE: British Society of Echocardiography
CABG: coronary artery bypass graft
CI: confidence interval
CMR: cardiovascular magnetic resonance
COVID-19: coronavirus disease 2019
CT: computed tomography
DD: diastolic dysfunction
DMD: Duchenne Muscular Dystrophy
EACVI: European Association of Cardiovascular Imaging
ED: emergency department
GLS: global longitudinal strain
HCM: hypertrophic cardiomyopathy
HFpEF: heart failure with preserved ejection fraction
HFrEF: heart failure with reduced ejection fraction
HTN: hypertension
HR: hazard ratio
IHD: ischemic heart disease
LAS: left atrial strain
LFLG: low-flow low-gradient
LGE: late gadolinium enhancement
LOA: limits of agreement
LV: left ventricle
LVEF: left ventricular ejection fraction
LVH: left ventricular hypertrophy
MACE: major adverse cardiovascular event
MAPSE: mitral annular plane systolic excursion
NYHA: New York Heart Association
OR: odds ratio
PAH: pulmonary artery hypertension
PE: pulmonary embolism
POCUS: point-of-care ultrasound
TAVI: Transaortic catheter valve implantation
TDI: tissue Doppler imaging
TMAD: tissue mitral annular displacement
TTE: transthoracic echocardiography
TEE: transesophageal echocardiography
RAVI: right atrial volume index
RV: right ventricle
SCMR: Society for Cardiovascular Magnetic Resonance
STE: speckle tracking echocardiography.

## Central graphical abstract

Clinical utility of MAPSE

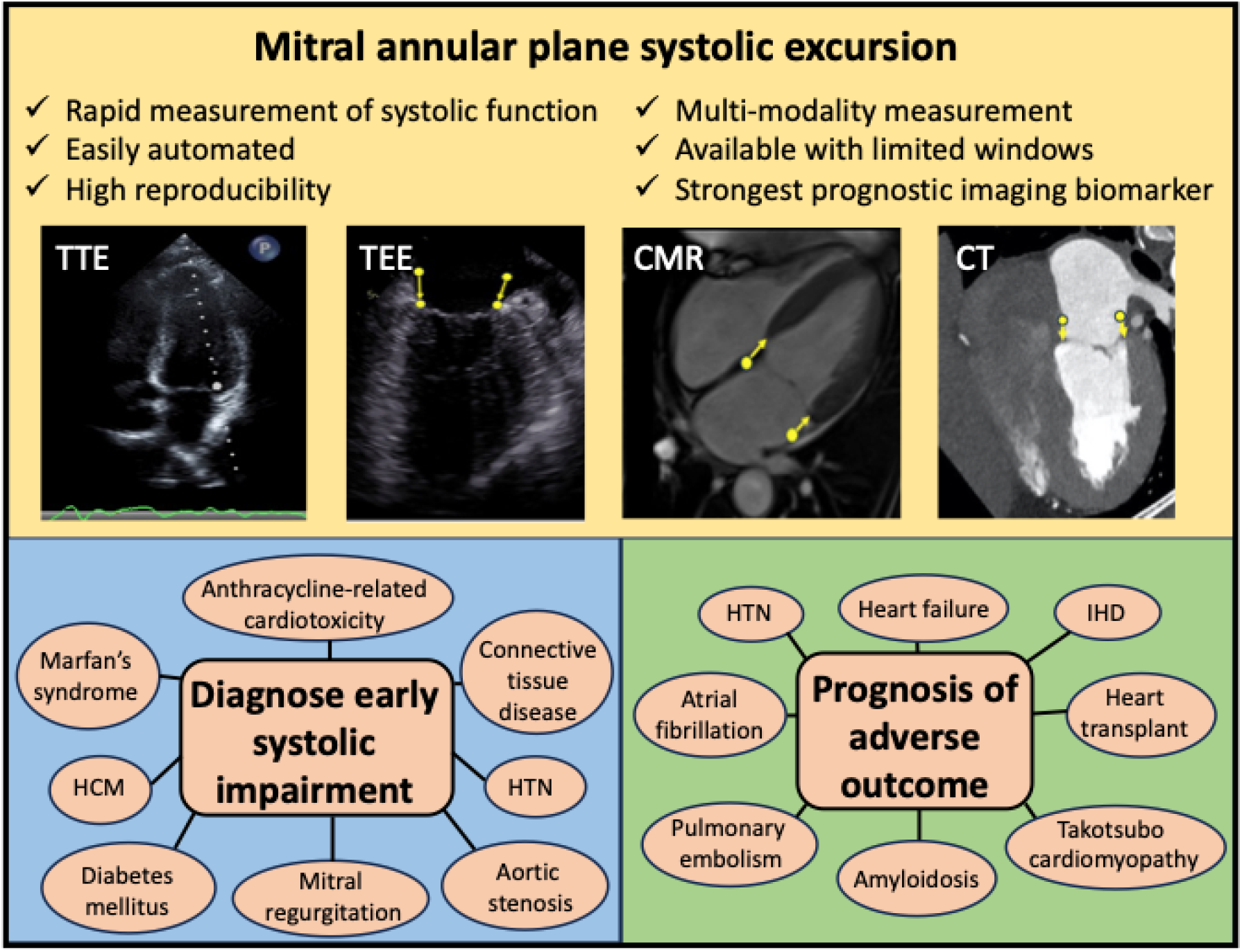

Central graphic created by authors (DF), individual panels reproduced and labelled with permission from Mayr^89^ (CMR), Yu^117^ (TOE) and Lorenzatti^118^ (CT).

## References

1. Keele KD. Leonardo da Vinci’s Corpus of Anatomical Studies. Hist Med. 1980;8:4–5.

2. Zaky A., Grabhorn L., Feigenbaum H. Movement of the mitral ring: a study in ultrasoundcardiography. Cardiovasc Res. 1967;1:121–31.

3. Carlsson M., Ugander M., Heiberg E., Arheden H. The quantitative relationship between longitudinal and radial function in left, right, and total heart pumping in humans. Am J Physiol Heart Circ Physiol. 2007;293:H636–44.

4. Carlsson M., Ugander M., Mosen H. et al. Atrioventricular plane displacement is the major contributor to left ventricular pumping in healthy adults, athletes, and patients with dilated cardiomyopathy. Am J Physiol Heart Circ Physiol. 2007;292:H1452–9.

5. Asgeirsson D,. Hedstrom E., Jogi J. et al. Longitudinal shortening remains the principal component of left ventricular pumping in patients with chronic myocardial infarction even when the absolute atrioventricular plane displacement is decreased. BMC Cardiovasc Disord. 2017;17:208.

6. Robinson S., Rana B., Oxborough D et al. A practical guideline for performing a comprehensive transthoracic echocardiogram in adults: the British Society of Echocardiography minimum dataset. Echo Res Pract. 2020;7:G59–G93.

7. Mitchell C., Rahko P.S., Blauwet L.A. et al. Guidelines for Performing a Comprehensive Transthoracic Echocardiographic Examination in Adults: Recommendations from the American Society of Echocardiography. J Am Soc Echocardiogr. 2019;32:1–64.

8. Galderisi M., Cosyns B., Edvardsen T. et al. Standardization of adult transthoracic echocardiography reporting in agreement with recent chamber quantification, diastolic function, and heart valve disease recommendations: an expert consensus document of the European Association of Cardiovascular Imaging. Eur Heart J Cardiovasc Imaging. 2017;18:1301–1310.

9. Kramer C.M., Barkhausen. J, Bucciarelli-Ducci C. et al. Standardized cardiovascular magnetic resonance imaging (CMR) protocols: 2020 update. J Cardiovasc Magn Reson. 2020;22:17.

10. Vancheri F., Longo G., Henein M.Y. Left ventricular ejection fraction: clinical, pathophysiological, and technical limitations. Front Cardiovasc Med. 2024;11:1340708.

11. Mizuguchi Y,. Oishi Y., Miyoshi H,. et al. The functional role of longitudinal, circumferential, and radial myocardial deformation for regulating the early impairment of left ventricular contraction and relaxation in patients with cardiovascular risk factors: a study with two-dimensional strain imaging. J Am Soc Echocardiogr. 2008;21:1138–44.

12. Slawinski G., Hawryszko M., Lizewska-Springer A. et al. Global Longitudinal Strain in Cardio-Oncology: A Review. Cancers (Basel). 2023;15:986.

13. Hu K., Liu D., Herrmann S. et al. Clinical implication of mitral annular plane systolic excursion for patients with cardiovascular disease. Eur Heart J Cardiovasc Imaging. 2013;14:205–12.

14. Doesch C., Sperb A., Sudarski S. et al. Mitral annular plane systolic excursion is an easy tool for fibrosis detection by late gadolinium enhancement cardiovascular magnetic resonance imaging in patients with hypertrophic cardiomyopathy. Arch Cardiovasc Dis. 2015;108:356–66.

15. Xue H., Artico J., Davies R.H. et al. Automated in-line artificial intelligence measured global longitudinal shortening and mitral annular plane systolic excursion: reproducibility and prognostic significance. J Am Heart Assoc. 2022;11:e023849.

16. Moher D., Liberati A., Tetzlaff J. et al. Preferred reporting items for systematic reviews and meta-analyses: the PRISMA statement. Ann Intern Med 2009;151:264–9.

17. Hunter J.E., Schmidt F.S. Methods of Meta-Analysis: Correcting Error and Bias in Research Findings. 2nd ed. Newbury Park, California.: Sage, 1990.

18. Wells GA, Shea B, O’Connell D, Peterson J, Welch V, Losos M, Tugwell P. The Newcastle-Ottawa Scale (NOS) for assessing the quality of nonrandomised studies in meta-analyses. Ottawa Hospital Research Institute. 2000. Available: http://www.ohri.ca/programs/clinical_epidemiology/oxford.asp

19. Herzog R, Álvarez-Pasquin MJ, Díaz C, Del Barrio JL, Estrada JM, Gil Á. Are healthcare workers’ intentions to vaccinate related to their knowledge, beliefs and attitudes? A systematic review. BMC Public Health. 2013;13:154.

20. Modesti PA, Reboldi G, Cappuccio FP, et al. Panethnic differences in blood pressure in Europe: a systematic review and meta-analysis. PLoS ONE. 2016;11(1):e0147601.

21. Carlhall C.J., Lindstrom L, Wranne B., Nylander E. Atrioventricular plane displacement correlates closely to circulatory dimensions but not to ejection fraction in normal young subjects. Clin Physiol. 2001;21:621–8.

22. Wang Y.H., Sun L., Li S.W. et al. Normal reference values for mitral annular plane systolic excursion by motion-mode and speckle tracking echocardiography: a prospective, multicentre, population-based study. Eur Heart J Cardiovasc Imaging. 2023;24:1384–1393.

23. Grue J.F., Storve S., Stoylen A. et al. Normal ranges for automatic measurements of tissue Doppler indices of mitral annular motion by echocardiography. Data from the HUNT3 Study. Echocardiography. 2019;36:1646–1655.

24. Ochs M.M., Fritz T., Andre F. et al. A comprehensive analysis of cardiac valve plane displacement in healthy adults: age-stratified normal values by cardiac magnetic resonance. Int J Cardiovasc Imaging. 2017;33:721–729.

25. Leng S., Zhao X,. Koh A.S. et al. Age-related changes in four-dimensional CMR-derived atrioventricular junction velocities and displacements: Implications for the identification of altered annular dynamics for ventricular function assessment. Int J Cardiol Heart Vasc .2019;22:6–12.

26. Simonson J.S., Schiller N.B. Descent of the base of the left ventricle: an echocardiographic index of left ventricular function. J Am Soc Echocardiogr. 1989;2:25–35.

27. Qin J.X., Shiota T., Tsujino H. et al. Mitral annular motion as a surrogate for left ventricular ejection fraction: real-time three-dimensional echocardiography and magnetic resonance imaging studies. Eur J Echocardiogr. 2004;5:407–15.

28. Elnoamany M.F., Abdelhameed A.K. Mitral annular motion as a surrogate for left ventricular function: correlation with brain natriuretic peptide levels. Eur J Echocardiogr. 2006;7:187–98.

29. Tsang W., Ahmad H., Patel A.R. et al. Rapid estimation of left ventricular function using echocardiographic speckle-tracking of mitral annular displacement. J Am Soc Echocardiogr. 2010;23:511–5.

30. Bazaz R., Edelman K., Gulyasy B., Lopez-Candales A. Evidence of robust coupling of atrioventricular mechanical function of the right side of the heart: insights from M-mode analysis of annular motion. Echocardiography. 2008;25:557–61.

31. Alam M. The atrioventricular plane displacement as a means of evaluating left ventricular systolic function in acute myocardial infarction. Clin Cardiol. 1991;14:588–94.

32. Alam M, Hoglund C, Thorstrand C. Longitudinal systolic shortening of the left ventricle: an echocardiographic study in subjects with and without preserved global function. Clin Physiol 1992;12:443–52.

33. Berg J., Akesson J., Jablonowski R. et al. Ventricular longitudinal function by cardiovascular magnetic resonance predicts cardiovascular morbidity in HFrEF patients. ESC Heart Fail. 2022;9:2313–2324.

34. Turan O., Kocabas A. Disturbed atrial conduction in patients with Duchenne Muscular Dystrophy. Anatol J Cardiol. 2024;28:493–8.

35. Zidan D.H., Helmy T.A. Usefulness of mitral annular plane systolic excursion in assessment of left ventricular systolic function in mechanically ventilated obese patients. J Crit Care. 2016;34:74–6.

36. Shah A., Nanjayya V., Ihle J. Mitral annular plane systolic excursion as a predictor of left ventricular ejection fraction in mechanically ventilated patients. Australas J Ultrasound Med. 2019;22:138–142.

37. Bergenzaun L., Ohlin H., Gudmundsson P. et al. Mitral annular plane systolic excursion (MAPSE) in shock: a valuable echocardiographic parameter in intensive care patients. Cardiovasc Ultrasound. 2013;11:16.

38. Brault C, Zerbib Y, Mercado P et al. Mitral annular plane systolic excursion for assessing left ventricular systolic dysfunction in patients with septic shock. BJA Open 2023;7:100220.

39. Matos J., Kronzon I., Panagopoulos G., Perk G. Mitral annular plane systolic excursion as a surrogate for left ventricular ejection fraction. J Am Soc Echocardiogr. 2012;25:969–74.

40. Lopez-Candales A., Hernandez-Suarez D.F., Lopez Menendez F. Mitral annular dynamics and left ventricular diastole. Cardiol Res. 2017;8:228–231.

41. Hernandez-Burgos P.M., Lopez-Menedez F., Candales M.D., Lopez-Candales A. Is mitral annular ascent useful in studying left ventricular function through left atrio-ventricular interactions? Indian Heart J. 2018;70:368–372.

42. Stenberg Y, Wallinder L, Lindberg A, et al. Preoperative point-of-care Aasessment of left ventricular systolic dysfunction With Ttansthoracic echocardiography. Anesth Analg. 2021;132:717–725.

43. Ozer P.K., Govdeli E.A., Demirtakan Z.G. et al. The relation of echo-derived lateral MAPSE to left heart functions and biochemical markers in patients with preserved ejection fraction: Short-term prognostic implications. J Clin Ultrasound. 2022;50:593–600.

44. Magdy G., Hamdy E., Elzawawy T., Ragab M. Value of mitral annular plane systolic excursion in the assessment of contractile reserve in patients with ischemic cardiomyopathy before cardiac revascularization. Indian Heart J. 2018;70:373–378.

45. Cirin L., Luca CT., Vacarescu C., et al. Added value of MAPSE to assess LV systolic function in conventional cardiac pacing. J Clin Med. 2025;14:6880.

46. Ballo P., Bocelli A., Motto A., Mondillo S. Concordance between M-mode, pulsed tissue Doppler, and colour tissue Doppler in the assessment of mitral annulus systolic excursion in normal subjects. Eur J Echocardiogr. 2008;9:748–53.

47. Mondillo S., Galderisi M., Ballo P. et al. Left ventricular systolic longitudinal function: comparison among simple M-mode, pulsed, and M-mode color tissue Doppler of mitral annulus in healthy individuals. J Am Soc Echocardiogr 2006;19:1085–91.

48. Liu L., Tuo S., Zhang J. et al. Reduction of left ventricular longitudinal global and segmental systolic functions in patients with hypertrophic cardiomyopathy: Study of two-dimensional tissue motion annular displacement. Exp Ther Med. 2014;7:1457–1464.

49. Chiu D.Y., Abidin N,. Hughes J., et al. Speckle tracking determination of mitral tissue annular displacement: comparison with strain and ejection fraction, and association with outcomes in haemodialysis patients. Int J Cardiovasc Imaging. 2016;32:1511–8.

50. Blixt P.J., Chew M.S., Ahman R et al. Left ventricular longitudinal wall fractional shortening accurately predicts longitudinal strain in critically ill patients with septic shock. Ann Intensive Care. 2021;11:52.

51. Iwakura K, Onishi T, Okamura A, et al. Tissue Mitral Annular Displacement in Patients With Myocardial Infarction — Comparison With Global Longitudinal Strain. Circ Rep. 2021;3(9):530–539. doi:10.1253/circrep.CR-21-0076

52. Ji X,. Zhang Y., Xie Y. et al. Feasibility and prognostic value of tissue motion annular displacement in patients with heart transplantation. Echocardiography. 2024;41:e15809.

53. Wenzelburger F.W., Tan Y.T,. Choudhary F.J., et al. Mitral annular plane systolic excursion on exercise: a simple diagnostic tool for heart failure with preserved ejection fraction. Eur J Heart Fail. 2011;13:953–60.

54. Teraguchi I., Hozumi T,. Takemoto K. et al. Assessment of decreased left ventricular longitudinal deformation in asymptomatic patients with organic mitral regurgitation and preserved ejection fraction using tissue-tracking mitral annular displacement by speckle-tracking echocardiography. Echocardiography. 2019;36:678–686.

55. Luszczak J., Olszowska M., Drapisz S. et al. Assessment of left ventricle function in aortic stenosis: mitral annular plane systolic excursion is not inferior to speckle tracking echocardiography derived global longitudinal peak strain. Cardiovasc Ultrasound. 2013;11:45.

56. Lopez-Candales A., Hernandez-Suarez D.F., Menendez F.L. Are measures of left ventricular longitudinal shortening affected by left atrial enlargement? Cardiol Res. 2018;9:1–6.

57. Zhang W., Azibani F,. Libhaber E. et al. The role of conventional echocardiographic parameters on detecting subclinical anthracycline therapy related cardiac dysfunction-The SATRACD study. Front Cardiovasc Med. 2022;9:966230.

58. Nemes A, Kalapos A, Domsik P, Forster T. Longitudinal systolic excursion of the mitral annular plane and left ventricular rotational mechanics are associated in healthy adults: three-dimensional speckle-tracking echocardiography-derived insights from the MAGYAR-Healthy Study. Journal of Ultrasound in Medicine. 2014;33(11):1861– 1866.

59. Xiao H.B., Kaleem S., McCarthy C., Rosen S.D. Abnormal regional left ventricular mechanics in treated hypertensive patients with ‘normal left ventricular function’. Int J Cardiol. 2006;112:316–21.

60. Koulouris S.N., Kostopoulos K.G., Triantafyllou K.A. et al. Impaired systolic dysfunction of left ventricular longitudinal fibers: a sign of early hypertensive cardiomyopathy. Clin Cardiol. 2005;28:282–6.

61. Ballo P., Cameli M., Mondillo S. et al. Impact of diabetes and hypertension on left ventricular longitudinal systolic function. Diabetes Res Clin Pract. 2010;90:209–15.

62. Salas-Pacheco J.L., Lomeli-Sanchez O., Baltazar-Gonzalez O., Soto M.E. Longitudinal systolic dysfunction in hypertensive cardiomyopathy with normal ejection fraction. Echocardiography. 2022;39:46–53.

63. Li Y., Wu C., Li Y. Feasibility study of automated cardiac motion quantification to assess left ventricular function in type 2 diabetes. Sci Rep. 2023;13:1101.

64. Loncarevic B., Trifunovic D,. Soldatovic I., Vujisic-Tesic B. Silent diabetic cardiomyopathy in everyday practice: a clinical and echocardiographic study. BMC Cardiovasc Disord. 2016;16:242.

65. Tromba L, Tartaglia F, Carbotta S et al. The role of sleeve gastrectomy in reducing cardiovascular Risk. Obes Surg. 2017;27:1145–1151.

66. Rydberg E., Gudmundsson P., Kennedy L., Erhardt L., Willenheimer R. Left atrioventricular plane displacement but not left ventricular ejection fraction is influenced by the degree of aortic stenosis. Heart. 2004;90:1151–5.

67. Takeda S., Rimington H., Smeeton N., Chambers J. Long axis excursion in aortic stenosis. Heart 2001;86:52–6.

68. Weidemann F, Herrmann S, Stork S et al. Impact of myocardial fibrosis in patients with symptomatic severe aortic stenosis. Circulation 2009;120:577–84.

69. Kempny A., Diller G.P., Kaleschke G. et al. Longitudinal left ventricular 2D strain is superior to ejection fraction in predicting myocardial recovery and symptomatic improvement after aortic valve implantation. Int J Cardiol. 2013;167:2239–43.

70. Matos J.D., Kiss J.E., Locke A.H., et al. Relation of the mitral annular plane systolic excursion to risk for intervention in initially asymptomatic patients with aortic stenosis and preserved systolic function. Am J Cardiol. 2017;120:2031–2034.

71. Herrmann S., Stork S., Niemann M. et al. Low-gradient aortic valve stenosis myocardial fibrosis and its influence on function and outcome. J Am Coll Cardiol. 2011;58:402–12.

72. Liu D., Hu K., Liebner E. et al. Value of low-dose dobutamine stress echocardiography on defining true severe low gradient aortic stenosis in patients with preserved left ventricular ejection fraction. Int J Cardiovasc Imaging 2018;34:1877–1887.

73. Dursunoglu D., Polat B., Evrengul H. et al. Assessment of systolic function by atrioventricular plane displacement in patients with diastolic dysfunction. Acta Cardiol. 2004;59:409–15.

74. Gromadzinski L., Pruszczyk P. Echocardiographic changes in patients with stage 3-5 chronic kidney disease and left ventricular diastolic dysfunction. Cardiorenal Med. 2014;4:234–43.

75. Taşolar H., Mete T., Cetin M. et al. Mitral annular plane systolic excursion in the assessment of left ventricular diastolic dysfunction in obese adults. Anatol J Cardiol. 2015;15:558–64.

76. Bytyci I., Bajraktari G. Left atrial changes in early stages of heart failure with preserved ejection fraction. Echocardiography. 2016;33:1479–1487.

77. Lopez-Candales A. Automated functional imaging for assessment of left ventricular mechanics in the presence of left ventricular hypertrophy. Echocardiography. 2014;31:605–14.

78. Wierzbowska-Drabik K., Chrzanowski L., Kapusta A. et al. Severe obesity impairs systolic and diastolic heart function - the significance of pulsed tissue Doppler, strain, and strain rate parameters. Echocardiography. 2013;30:904–11.

79. Qureshi A.E., Chaudhary F.U.. Mitral annular plan systolic excursion (MAPSE) underestimates left ventricular systolic function in patients with left ventricular hypertrophy. The Professional Medical Journal 2018;25:1622–6.

80. Fröjdh F., Soundappan D., Sörensson P., et al. Strain measures of the left ventricle and left atrium are composite measures of left heart geometry and function. MedRxiv (Preprint) May 2025.

81. Ozden O., Bingol G., Unlu S., Boyuk F. A cardiac magnetic resonance study: comparison of biventricular longitudinal function in hypertrophic cardiomyopathy patients and normal individuals. Cureus. 2023;15:e34165.

82. Kassab J., Hajj J., Harb R., et al. Mitral annulat function in mitral annular calcification and severe regurgitation. JACC Advances. 2025;4:102161.

83. Kiotsekoglou A., Bajpai A., Bijnens B.H. et al. Early impairment of left ventricular long-axis systolic function demonstrated by reduced atrioventricular plane displacement in patients with Marfan syndrome. Eur J Echocardiogr. 2008;9:605–13.

84. Harries I., Biglino G., Ford K. et al. Prospective multiparametric CMR characterization and MicroRNA profiling of anthracycline cardiotoxicity: A pilot translational study. Int J Cardiol Heart Vasc. 2022;43:101134.

85. Ichikawa N, Nishizaki Y, Miyazaki S et al. Efficacy of mitral annular velocity as an alternative marker of left ventricular global longitudinal strain to detect the risk of cancer therapy-related cardiac disorders. Echocardiography 2024;41:e15877.

86. Botelho L.F.B., de Melo M.D.T., de Almeida A.L.C., Salemi V.M.C. Accuracy of mitral annular plane systolic excursion in diagnosing anthracycline-induced subclinical cardiotoxicity in patients with breast cancer - a retrospective cohort study. Cardiooncology. 2024;10:76.

87. Tafarshiku R., Henein M.Y., Berisha-Muharremi V. et al. Left ventricular diastolic and systolic functions in patients with hypothyroidism. Medicina (Kaunas). 2020;56:524.

88. Berger S.G., Witczak B.N., Reiseter S. et al. Cardiac dysfunction in mixed connective tissue disease: a nationwide observational study. Rheumatol Int. 2023;43:1055–1065.

89. Rangarajan V., Chacko S.J., Romano S .et al. Left ventricular long axis function assessed during cine-cardiovascular magnetic resonance is an independent predictor of adverse cardiac events. J Cardiovasc Magn Reson. 2016;18:35.

90. Romano S., Judd R.M., Kim R.J. et al. Left ventricular long-axis function assessed with cardiac cine MR imaging is an independent predictor of all-cause mortality in patients with reduced ejection fraction: a multicenter study. Radiology. 2018;286:452–460.

91. Svealv B.G., Olofsson E.L., Andersson B. Ventricular long-axis function is of major importance for long-term survival in patients with heart failure. Heart. 2008;94:284–9.

92. Willenheimer R., Cline C., Erhardt L., Israelsson B.. Left ventricular atrioventricular plane displacement: an echocardiographic technique for rapid assessment of prognosis in heart failure. Heart. 1997;78:230–6.

93. Tavazzi G., Colombo C.N.J. Klersy C. et al. Echocardiographic parameters for weaning from Extracorporeal Membrane Oxygenation - the role of longitudinal function and cardiac time intervals. Eur Heart J Cardiovasc Imaging. 2024;26:359–67.

94. Berg J., Jablonowski R., Mohammad M., et al. Ventricular longitudinal shortening is an independent predictor of death in heart failure patients with reduced ejection fraction. Sci Rep. 2021;11:20280.

95. Mayr A., Pamminger M., Reindl M. et al. Mitral annular plane systolic excursion by cardiac MR is an easy tool for optimized prognosis assessment in ST-elevation myocardial infarction. Eur Radiol. 2020;30:620–629.

96. Wang L., Yuan W., Huang X., Zhao X., Zhao X.. Cardiac magnetic resonance-derived mitral annular plane systolic excursion: a robust indicator for risk stratification after myocardial infarction. Int J Cardiovasc Imaging. 2024;40:897–906.

97. Brand B., Rydberg E., Ericsson G., Gudmundsson P., Willenheimer R. Prognostication and risk stratification by assessment of left atrioventricular plane displacement in patients with myocardial infarction. Int J Cardiol 2002;83:35–41.

98. Yan C., Chang Y., Fang-Wu et al. Evaluation of the prognostic value of lateral MAPSE in patients with suspected coronary artery disease. Int J Cardiol Heart Vasc. 2025;56:101567.

99. Holzknecht M., Reindl M., Tiller C. et al. Global longitudinal strain improves risk assessment after ST-segment elevation myocardial infarction: a comparative prognostic evaluation of left ventricular functional parameters. Clin Res Cardiol. 2021;110:1599–1611.

100. Rydberg E., Erhardt L., Brand B., Willenheimer R. Left atrioventricular plane displacement determined by echocardiography: a clinically useful, independent predictor of mortality in patients with stable coronary artery disease. J Intern Med. 2003;254:479–85.

101. El-Battrawy I., Ansari U., Lang S. et al. Risk stratification in Takotsubo syndrome: a role of mitral annular plane systolic excursion. QJM 2018;111:231–236.

102. Romano S., Judd R.M., Kim R.J. et al. Prognostic Implications of Mitral Annular Plane Systolic Excursion in patients with hypertension and a clinical indication for cardiac magnetic resonance imaging: a multicenter study. JACC Cardiovasc Imaging. 2019;12:1769–1779.

103. Ballo P., Barone D., Bocelli A., Motto A., Mondillo S. Left ventricular longitudinal systolic dysfunction is an independent marker of cardiovascular risk in patients with hypertension. Am J Hypertens 2008;21:1047–54.

104. Siepen F.A.D., Bauer R., Voss A. et al. Predictors of survival stratification in patients with wild-type cardiac amyloidosis. Clin Res Cardiol. 2018;107:158–169.

105. Ochs M.M., Fritz T., Arenja N. et al. Regional differences in prognostic value of cardiac valve plane displacement in systemic light-chain amyloidosis. J Cardiovasc Magn. Reson 2017;19:87.

106. Riffel J.H., Mereles D., Emami M. et al. Prognostic significance of semiautomatic quantification of left ventricular long axis shortening in systemic light-chain amyloidosis. Amyloid. 2015;22:45–53.

107. Liu D., Hu K., Stork S. et al. Predictive value of assessing diastolic strain rate on survival in cardiac amyloidosis patients with preserved ejection fraction. PLoS One. 2014;9:e115910.

108. Lindholm A., Kjellstrom B., Seemann F. et al. Atrioventricular plane displacement and regional function to predict outcome in pulmonary arterial hypertension. Int J Cardiovasc Imaging. 2022;38:2235–2248.

109. Havaldar A.A.. Evaluation of sepsis induced cardiac dysfunction as a predictor of mortality. Cardiovasc Ultrasound. 2018;16:31.

110. Rydberg E., Arlbrandt M., Gudmundsson P., et al. Left atrioventricular plane displacement predicts cardiac mortality in patients with chronic atrial fibrillation. Int J Cardiol 2003;91:1–7.

111. Matos J.D. Balachandran I., Heidinger B.H. et al. Mitral annular plane systolic excursion and tricuspid annular plane systolic excursion for risk stratification of acute pulmonary embolism. Echocardiography. 2020;37:1008–1013.

112. Jarori U., Maatman T.K., Maatman B., et al. Mitral annular plane systolic excursion: an early marker of mortality in severe COVID-19. J Am Soc Echocardiogr. 2020;33:1411–1413.

113. Vahanian A., Beyersdorf F., Praz F. et al. 2021 ESC/EACTS Guidelines for the management of valvular heart disease. Eur Heart J. 2022;43:561–632.

114. Fukui M., Hashimoto G., Lopes B.B.C. et al. Association of baseline and change in global longitudinal strain by computed tomography with post-transcatheter aortic valve replacement outcomes. Eur Heart J Cardiovasc Imaging. 2022;23:476–484.

115. Grue J.F., Storve S., Dalen H. et al. Automatic measurements of mitral annular plane systolic excursion and velocities to detect left ventricular dysfunction. Ultrasound Med Biol. 2018;44:168–176.

116. Stoylen A., Skjaerpe T. Systolic long axis function of the left ventricle. Global and regional information. Scand Cardiovasc J. 2003;37:253–8.

117. Goodreads. Leonardo da Vinci > Quotes > Quotable Quote. March 1st 2025. https://www.goodreads.com/quotes/9010638-simplicity-is-the-ultimate-sophistication-when-once-you-have-tasted.

