## Supplementary methods for "The diagnostic and prognostic utility of mitral annular plane systolic excursion (MAPSE): A systematic review"

Supplemental methods

Search string:

PUBMED All 1946- June 15^th^ 2025

1. MAPSE
2. Mitral annulus planar systolic excursion
3. AVPD
4. Atrioventricular plane displacement
5. Mitral ring movement
6. Systolic impairment
7. Prognosis
8. Monitoring
9. 1 or 2 or 3 or 4 or 5
10. 6 or 7 or 8
11. 9 and 10

**Table S-1.** Excluded studies from those assessed for eligibility

| Author (year) | Reason for exclusion | DOI/ link |
| --- | --- | --- |
| Kumar (2025) | Abstract only | 10.5005/jaypee-journals-10071-24933.18 |
| Swaminathan (2025) | Abstract only | 10.5005/jaypee-journals-10071-24933.121 |
| Djermane (2025) | Abstract only | [10.1016/j.acvd.2024.10.112](https://doi.org/10.1016/j.acvd.2024.10.112) |
| Yin (2017) | No methods described for measuring MAPSE | 10.1371/journal.pone.0182881 |
| Blixt (2024) | Incorrect outcome measure | 10.1186/s13613-023-01235-5 |
| Zou (2020) | No methods described for measuring MAPSE | 10.1155/2020/5175393 |
| Stoylen (2018) | Dataset presented in other study | 10.1111/echo.13825 |
| Ma (2022) | Incorrect outcome measure | 10.1111/echo.15308 |
| Strzelczyk (2021) | Incorrect outcome measure | 10.1007/s11695-021-05710-5 |
| Stoylen (2020) | Incorrect outcome measure | 10.1136/openhrt-2020-001243 |
| Doesch (2016) | Dataset presented in other study | 10.7150/ijms.13530 |
| Bytici (2019) | Incorrect outcome measure | 10.1111/echo.14377 |
| Batalli (2017) | Incorrect outcome measure | 10.1186/s12947-017-0103-x |
| Lakkas (2020) | Incorrect outcome measure | 10.1111/echo.14570 |
| Jiang (2023) | Incorrect outcome measure | 10.1111/jog.15699 |
| Baron (2016) | Incorrect outcome measure | 10.1093/ehjci/jev160 |
| Cirin (2024) | Datasets presented in other study (review article) | 0.3390/jcm13175265 |
| Arenja (2017) | Incorrect outcome measure | 0.1007/s12410-017-9429-x |
| Ehrman (2018) | Datasets presented in other study (review article) | 10.1186/s13054-018-2043-8 |
| Giovanardi (2020) | Incorrect outcome measure | 10.3390/jcdd7010004 |
| Alatic (2022) | Incorrect outcome measure | 10.1155/2022/2746304 |
| Madry (2019) | Incorrect outcome measure | 10.15557/JoU.2019.0002 |
| Al-Amir (2024) | Incorrect outcome measure | 10.62877/9-IJCBS-24-25-19-9 |
| Zhang (2023) | Incorrect outcome measure | 10.1186/s12871-023-02142-9 |
| Tiller (2022) | Incorrect outcome measure | 10.1093/eurheartj/ehac544.266 |
| Külahcioglu  (2021) | Incorrect outcome measure | 10.5798/dicletip.1037638 |
| Prastaro (2017) | Datasets presented in other study (review article) | 10.1016/j.echo.2017.01.020 |
| Arenja (2017) | Datasets presented in other study (review article) | 10.1007/s12410-017-9429-x |
| Tasken (2024) | Incorrect outcome measure | 10.1016/j.ultrasmedbio.2024.01.017 |
| Moderato (2020) | Abstract only | 10.1093/ehjci/jez319.976 |
| Yu (2025) | Doctoral thesis | https://ntnuopen.ntnu.no/ntnu-xmlui/handle/11250/3174251 |
| Schick (2023) | Incorrect outcome measure | [10.1016/j.ajem.2023.03.018](https://doi.org/10.1016/j.ajem.2023.03.018) |
| Koseoglu (2024) | Incorrect outcome measure | 10.4149/BLL_2024_78 |
| Spalla (2018) | Incorrect outcome measure | 10.1016/j.jvc.2018.04.005 |
| Matos (2014) | Abstract only | 10.1016/j.amjcard.2017.08.021 |
| Baron (2015) | Abstract only | 10.1093/ehjci/jev268 |
| Nordbeck (2021) | Abstract only | 10.1161/circ.144.suppl_1.137 |
| Palppert (2019) | Abstract only | 10.1093/ehjci/jez122.004 |
| Von Jeinsen (2018) | Abstract only | www.ahajournals.org/doi/abs/10.1161/circ.138.suppl_1.12771 |

**Table S-2**. Newcastle-Ottowa scale for cohort studies.

| **Domain** | **Item** | **Maximum score** | **Potential reasons for a reduction in score** |
| --- | --- | --- | --- |
| Selection | Representativesness of cohort | 1 | Unclear how participants recruited, or selected from specialised group when intended to represent wider population |
|  | Selection of non-exposed cohort | 1 | No comparison group or no internal control |
|  | Ascertainment of exposure | 1 | Exposure not measured using validated or reproducible method, or method not described |
|  | Demonstration that outcome not present at the start | 1 | Baseline status unclear, or timing between exposure and outcome not stated |
| Comparability | Comparability of cohorts on basis of design or analysis | 2 | No adjustment for confounding factors, or incomplete multivariable modelling |
| Outcome | Assessment of outcome | 1 | Outcome not clearly defined, or based on self reporting only |
|  | Follow up duration | 1 | Follow up duration too short for chosen outcomes |
|  | Adequacy of follow up | 1 | Significant or unreported loss to follow up, or no description of follow up |

**Table S-3**. Modified Newcastle-Ottowa Scale for cross-sectional studies.

| **Domain** | **Item** | **Maximum score** | **Potential reasons for a reduction in score** |
| --- | --- | --- | --- |
| Selection | Representativeness of cohort | 1 | Not representative of target population |
|  | Sample size justification | 1 | No justification for size |
|  | Reporting of those not recruited | 1 | No description of reasons for exlusion/ numbers excluded |
|  | Ascertainment of the exposure | 2 | Use of non-validated measurement technique, or technique not clearly described |
| Comparability | Comparability of groups on basis of design or analysis | 2 | Confounding factors not controlled (or minimally controlled) |
| Outcome | Assessment of outcome | 2 | Outcomes self reported or assessment not described |
|  | Statistical test | 1 | Not described or not appropriate |

**Table S-4.** Modified Newcastle-Ottowa Scale for *cross-sectional* studies comparing MAPSE values when ejection fraction is normal.

| Author (year) | Selection  (/5) | Comparability  (/2) | Outcome  (/3) | Total score (/10) | Quality rating |
| --- | --- | --- | --- | --- | --- |
| Xiao (2006) | 3 | 1 | 3 | 7 | High |
| Koulouris (2005) | 3 | 1 | 3 | 7 | High |
| Ballo (2010) | 3 | 2 | 3 | 8 | High |
| Salas-Pachecho (2021) | 3 | 1 | 3 | 7 | High |
| Li (2023) | 3 | 1 | 3 | 7 | High |
| Loncarevic (2016) | 3 | 1 | 3 | 7 | High |
| Takeda (2001) | 3 | 1 | 3 | 7 | High |
| Rydberg (2004) | 3 | 2 | 3 | 8 | High |
| Dursunoglu (2004) | 3 | 1 | 3 | 7 | High |
| Gromadzinski (2014) | 3 | 1 | 3 | 7 | High |
| Wenzelburger (2011) | 4 | 1 | 3 | 8 | High |
| Tasolar (2015) | 5 | 1 | 3 | 9 | High |
| Bytyci (2016) | 3 | 2 | 3 | 8 | High |
| Wierbowska-Drabik (2013) | 4 | 1 | 3 | 8 | High |
| Lopez-Candales (2014) | 4 | 2 | 3 | 9 | High |
| Qureshi (2018) | 3 | 1 | 3 | 7 | High |
| Doesch (2015) | 3 | 1 | 3 | 7 | High |
| Ozden (2023) | 4 | 1 | 3 | 8 | High |
| Kiotsekoglou (2008) | 3 | 2 | 3 | 8 | High |
| Tafarshiku (2020) | 3 | 1 | 3 | 7 | High |
| Berger (2023) | 4 | 1 | 3 | 8 | High |
| Kassab (2025) | 4 | 1 | 3 | 8 | High |

| Author (year) | Design | Selection (/4) | Comparability (/2) | Outcome (/3) | Total score (/9) | Quality rating |
| --- | --- | --- | --- | --- | --- | --- |
| Tromba (2017) | Prospective cohort | 3 | 0 | 3 | 6 | Moderate |
| Weidemann (2009) | Prospective cohort | 4 | 2 | 3 | 9 | High |
| Herrmann (2011) | Prospective cohort | 3 | 2 | 3 | 8 | High |
| Kempny (2013) | Prospective cohort | 4 | 1 | 3 | 8 | High |
| Matos (2017) | Retrospective cohort | 3 | 1 | 3 | 7 | High |
| Liu (2018) | Prospective cohort | 3 | 2 | 3 | 8 | High |
| Harries (2022) | Prospective cohort | 4 | 1 | 3 | 8 | High |
| Botelho (2024) | Retrospective cohort | 4 | 1 | 3 | 8 | High |
| Ichikawa (2024) | Retrospective cohort | 4 | 1 | 3 | 8 | High |

**Table S-5.** Newcastle-Ottowa Scale for *cohort* studies comparing MAPSE values when ejection fraction is normal.

**Table S-6**: Newcastle-Ottowa Scale for studies describing prognostic value of MAPSE.

| Author (year) | Design | Selection  (/4) | Comparability  (/2) | Outcome  (/3) | Total score (/9) | Quality rating |
| --- | --- | --- | --- | --- | --- | --- |
| Rangarajan (2016) | Prospective cohort | 4 | 2 | 3 | 9 | High |
| Ozer (2022) | Prospective cohort | 4 | 2 | 2 | 8 | High |
| Xue (2023) | Prospective cohort | 4 | 2 | 3 | 9 | High |
| Willenheimer (1997) | Prospective cohort | 4 | 2 | 3 | 9 | High |
| Svealve (2008) | Prospective cohort | 4 | 2 | 3 | 9 | High |
| Romano (2018) | Prospective cohort | 4 | 2 | 3 | 9 | High |
| Tavazzi (2024) | Prospective cohort | 4 | 1 | 3 | 8 | High |
| Berg (2021) | Prospective cohort | 4 | 2 | 3 | 9 | High |
| Brand (2002) | Prospective cohort | 4 | 2 | 3 | 9 | High |
| Rydberg (2003) | Prospective cohort | 4 | 2 | 3 | 9 | High |
| Mayr (2020) | Prospective cohort | 4 | 2 | 3 | 9 | High |
| Holzknecht (2021) | Prospective cohort | 4 | 2 | 3 | 9 | High |
| Iwakura (2021) | Prospective cohort | 4 | 2 | 3 | 9 | High |
| Wang (2024) | Retrospective cohort | 4 | 2 | 3 | 9 | High |
| Yan (2024) | Prospective cohort | 4 | 2 | 3 | 9 | High |
| El-Battrawy (2018) | Retrospective cohort | 4 | 2 | 2 | 8 | High |
| Romano (2018) | Retrospective cohort | 4 | 2 | 3 | 9 | High |
| Ballo (2008) | Prospective cohort | 4 | 2 | 3 | 9 | High |
| Dem Siepen (2018) | Retrospective cohort | 4 | 2 | 3 | 9 | High |
| Liu (2014) | Retrospective cohort | 3 | 2 | 3 | 8 | High |
| Riffel (2014) | Retrospective cohort | 4 | 2 | 3 | 9 | High |
| Ochs (2017) | Retrospective cohort | 4 | 2 | 2 | 8 | High |
| Ji (2024) | Prospective cohort | 4 | 2 | 3 | 9 | High |
| Lindholm (2022) | Retrospective cohort | 4 | 2 | 3 | 9 | High |
| Bergenzaun (2013) | Prospective cohort | 4 | 2 | 3 | 9 | High |
| Havaldar (2018) | Prospective cohort | 4 | 2 | 3 | 9 | High |
| Rydberg (2003) | Retrospective cohort | 4 | 2 | 3 | 9 | High |
| Matos (2020) | Retrospective cohort | 4 | 2 | 3 | 9 | High |
| Jarori (2020) | Retrospective cohort | 3 | 2 | 3 | 8 | High |
